# Implications of predator species richness in terms of zoonotic spillover transmission of filoviral hemorrhagic fevers in Africa

**DOI:** 10.1101/2023.02.12.23285832

**Authors:** Taehee Chang, Sung-il Cho, Kyung-Duk Min

## Abstract

Previous studies found that higher species richness of predators could reduce spillover risks of rodent-borne diseases. However, the effects on bat-borne diseases remains to be investigated. To this regard, we evaluated associations between predator species richness and the spillover events of *Ebolavirus* and *Marburgvirus*, the highly pathogenic bat-borne diseases in Africa. Stacked species distribution model approach was used to estimate predator species richness and Logistic regression analyses that considered spatiotemporal autocorrelations were conducted. The results showed that the third quartile (OR = 0.02, 95% CI 0.00–0.84) and fourth quartile (0.07, 0.00–0.42) of species richness of Strigiformes and the third quartile (0.15, CI 0.01–0.73) and fourth quartile (0.53, 0.03–0.85) of Colubridae showed significantly lower risks of spillover transmission of *Ebolavirus*. However, no significant association was found between predator species richness and *Marburgvirus* spillover. The results support a possible effect of predator species diversity on spillover suppression.

## Introduction

*Ebolavirus* and *Marburgvirus* are non-segmented, negative-stranded RNA viruses belonging to the family Filoviridae, a subgroup of the order Mononegavirales (1). There are six virus species in the *Ebolavirus* genus (Ebola virus, Sudan virus, Bombali virus, Tai Forest virus, Bundibugyo virus, and Reston virus) and two species in the *Marburgvirus* genus (Marburg virus and Ravn virus) (2). With the exception of the Reston virus, the viruses are considered indigenous to Africa, where multiple human outbreaks have occurred (3, 4). Filovirus epidemics cause catastrophic losses of human and animal life given the high case fatality rates, which are typically 60–70% but can reach 90% (1).

Significant progress in shortening the list of potential filovirus reservoir hosts has been made during the past decade. Apart from the *Rousettus aegypticus* fruit bat, which has repeatedly tested positive for *Marburgvirus*, antibodies against various Ebola species have been found in at least 14 other species of bats; however, only *Epomops franqueti, Hypsignathus monstrosus*, and *Myonycteris torquata* tested positive using PCR methods (5-8). The viruses may spread to other animals, including non-human primates, duikers (antelopes), or humans, from bat species shown to be vulnerable to filoviruses. Humans might contract the virus by handling or eating so-called bushmeat, such as roosting bats close to human dwellings, or via contact with infected mammalian bodily fluids (1).

Predators impact prey density, distribution, and behavior both directly and indirectly. Theoretically, such impacts might cascade to lower trophic levels and thus reduce the risk of zoonotic spillover (9, 10). Generalist predators (e.g., certain snakes, cats, owls, and raptors) that are either non-specialized in terms of prey selection and can thus move among target species, or that are highly mobile and therefore wander in search of better hunting grounds, have been suggested to chronically suppress prey numbers and thus stabilize population dynamics (9, 11). Predator non-lethal effects can influence the behavioral patterns of prey and reduce prey fitness. The predatory risk cues detected by prey, including visual, auditory, or chemical signals, allow them to identify the presence of predators and consequently alter their behavior in response to the danger of predation (12, 13). Few vertebrate predators specialize in hunting bats, and bat predation appears to be mainly opportunistic in nature. However, generalist and opportunistic predators may exert substantial effects on bat ecology, eventually reducing the rate of contact between reservoir hosts and humans and thus mitigating the risk of zoonotic spillover (9, 12, 14).

We hypothesized that high predator species richness will reduce the zoonotic spillover of filoviruses in Africa. We examined the associations between predator species richness and historical spillovers of *Ebolavirus* and *Marburgvirus* based on distributional data from known predators of bats only, as well as satellite-derived environmental data.

## Methods

### Study design and study area

In this ecological study, we used stacked species distribution models and the maximum entropy method (Maxent modeling) to calculate the number of predator species. We considered potential confounding factors when conducting logistic regression analyses of the relationship between predator species richness and spillover risk. We included all African countries with at least one reported human case of Ebola or Marburg infection. We confined the study regions to areas proposed in previous studies to harbor the reservoir species *E. franqueti, H. monstrosus*, and *M. torquata* of *Ebolavirus* and the *R. aegypticus* fruit bat of *Marburgvirus* (5-8). The distribution ranges were constructed using the geographical database of the International Union for the Conservation of Nature (IUCN) (15). To examine the relationship between predator species richness and filovirus cases, three datasets were compiled: (i) a comprehensive list of index case locations, (ii) geographical information on the distributions of predators and reservoir hosts, and (iii) environmental factors suggested to be ecologically significant. R software (v. 4. 2. 1) (16) was used for all data processing and analyses. The “dismo” package (17) was employed to model species niches. Bayesian parameter estimations that considered spatial and spatiotemporal autocorrelations were conducted using the “CARBayes” and “CARBayesST” packages (18).

### Outcome definitions

We identified index cases and rebuilt zoonotic spillover cases in both space and time. We searched the formal scientific literature using PubMed and the Web of Science for data on all historical filovirus outbreaks (3-8, 19). We sought to recreate the outbreaks in detail and locate the most likely index cases, thus infected humans who had interacted with disease-causing non-human sources. Cases reported between 2000 and 2021 were included in analysis because the environmental covariates used in the present report share their temporal ranges since that time. On the map of the study regions, we generated °× °grids and classified them in terms of their intersections with the point locations of index cases.

### Niche modeling and diversity maps

The suitability of habitats for natural predators of bats, i.e., the order Accipitriformes, Strigiformes, and Carnivora and family Colubridae and reservoir hosts in the order Chiroptera, was predicted using a simple species distribution modeling strategy (also termed ecological niche modeling), which integrates the reported occurrences of species with local climatic and geographic information. We used a maximum entropy approach (Maxent modeling) (20); this is one of the most widely used models when identifying species distributions. The approach employs presence-only data, which are helpful when modeling small and mobile species because it is (appropriately) challenging to establish their absence.

The occurrence data of included species within the study area (thus the African continent: 12.69 to – 22.42 N and 41.57 to –14.97 E) were those of the Global Biodiversity Information Facility. Species that occurred at more than 10 points were included in the models. Climatic and geographical data served as predictive variables when simulating the distributions of the species. The bioclimatic variables were derived from WorldClim ver. 2.0 (21) and the elevation data from the Shuttle Radar Topography Mission (ver. 4) (22) with a spatial resolution of 2.5 minutes (∼20 km^2^). The variables included in the modeling process were considered ecologically crucial in terms of species distribution, and they evidenced one-to-one intercorrelations < 0.7. These variables were the mean diurnal range (Bio02), temperature seasonality (Bio04), maximum temperature in the warmest month (Bio05), precipitation in the wettest quarter (Bio16), precipitation in the warmest quarter (Bio18), precipitation in the coldest quarter (Bio19), and the elevation. When fitting the models for each species, all variables were verified using the Jackknife test (20).

We used k-fold cross-validation to assess the models. Next, we converted the habitat suitability into a binary value (suitable habitat 1; unsuitable habitat 0). The threshold was the modeled prevalence closest to the observed prevalence. The numbers of species for which suitable habitat pixels in each grid exceeded 50% of the total grid areas were counted.

### Data acquisition and preprocessing

Global climatic data from 1970 to 2000 were collected from WorldClim ver. 2.0 (21), which features a spatial resolution of 2.5 minutes (∼20 km^2^) and offers monthly average precipitation and temperature data in raster format. The average annual precipitation and temperature for each grid region were computed.

The geographical confounding factors collected included elevation land cover, agricultural land use, and forest cover data. We obtained elevation data from the Shuttle Radar Topography Mission (22), which offers 90-m-scale worldwide elevation data in raster format. The values for each grid were averaged. Data on agricultural land use during 2000–2021 were gathered in raster format (23). The dataset contains the most likely International Geosphere-Biosphere Programme class for each 0.05ºpixel, and we calculated the proportion of each International Geosphere-Biosphere Programme class for all grids of interest. The Global Forest Change (GFC) (24) data yielded forest cover information. In terms of tree canopy cover, the likelihood of a tree canopy is presented in raster format and ranges from 0 to 100. We used 75 as the cutoff when determining whether a raster cell included a forest. We collected data on forest loss, defined as a change from a forest to a non-forest state, during 2000–2021. We then computed the proportion of forest coverage in each grid by subtracting the area of forest loss from that of the tree canopy cover.

The sociodemographic factors analyzed were the gross domestic product, human development index, population density, and human footprint score. Gross domestic product and human development index data from 1990–2015 were gathered, and the average values for each grid computed (25). The impact of human activity on the environment during 2000–2018 was measured using the human footprint score, which presents more significant anthropogenic pressures as higher scores (26). The values for each grid were averaged. Population density data were acquired from the WorldPop website (27). A population count dataset of the unconstrained global mosaics from 2000–2020 at a resolution of 1 km was used to calculate the population density for each grid.

Some variables did not cover the entire period from 2000 to 2021; in such cases, data from previous years were used to fill in for missing data. Detailed descriptions of each variable, including the temporal range and spatial resolution, can be found in (Supplementary Table 1).

### Statistical analysis

We developed logistic regression models for the study grids to determine odds ratios (ORs) with 95% confidence intervals (95% CIs) for the relationship between predator species richness and filovirus cases. We adjusted for all possible confounders except for variables with a Pearson correlation coefficient > 0.7 and variance inflation factor (VIF) > 10. To handle potential species richness overestimation, the indicators of predator species richness were entered into saturated models as categorical variables. We defined the Ebolavirus categories by quartiles and the Marburgvirus categories by medians. The categories with the lowest number of predator species served as the base categories.

We used Bayesian spatiotemporal models to derive the spatial and temporal patterns over 100,000 iterations with a burn-in of 95,000 when the model residuals were autocorrelated as revealed by the Moran I test and Durbin–Watson test. The following are the mathematical expressions of the models:

Model 1: 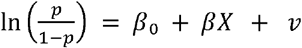

Model 2: 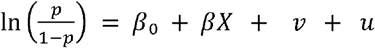

Model 3: 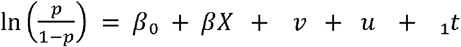

Model 4: 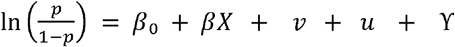

Model 5: 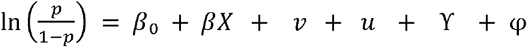

Where p is the probability of filovirus emergence; the constant β_0_ is the intercept; β_n_ is the regression coefficient; is the set of predictive variables; *v*_n_ is the non-spatial random component for grid n; *u*_n_ is the structured spatial random component for grid n; *t* is the temporal trend of the data with a constant term *a*_1_; φm random-walk component; and the random effect φ mn is the space–time interaction term. To choose the model affording the best performance in terms of the Bayesian framework, we compared Models 1–5 using the deviance information criterion (DIC) and the Watanabe–Akaike information criterion (WAIC).

### Sensitivity analysis

We performed sensitivity analysis using the “R-INLA” package for Bayesian parameter estimation (28). We estimated species richness using a geographical database from the IUCN website (15). Only species categorized as ‘extant’ that overlapped the species included in Maxent modeling were used. The numbers of species, the distribution ranges of which spanned more than 50% of a specific grid, were counted after intersecting the polygons representing the range data for the species. Finally, we constructed models using the species richness variables calculated via Maxent modeling and included only species reported to prey on bats.

## Results

### Processing datasets

In total, 32 index cases of *Ebolavirus* and 12 of *Marburgvirus* were identified across the African continent. The times of disease occurrence span the last four decades, starting with the first *Marburgvirus* case in 1976 (Figure 1A). The locations of the outbreaks spanned from Guinea in West Africa to Uganda and Kenya in East Africa (Figure 1B). We classified study grids by their intersections with the point locations of index cases reported between 2000 and 2021 (Figure 2A, B).

**Fig 1.**
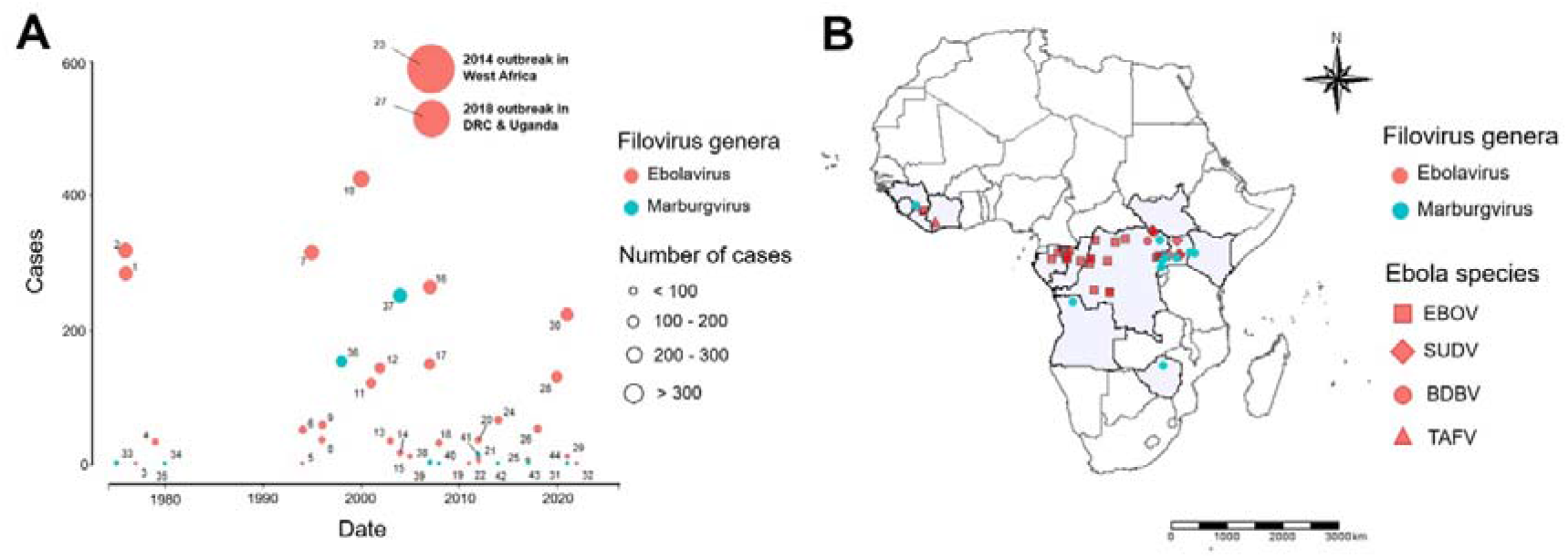
The locations and points of occurrence of filovirus outbreaks in Africa. (A) Shows the reported outbreaks of *Ebolavirus* and *Marburgvirus* through time, with its height along the y-axis reflect the number of cases. (B) Illustrates a map of the index cases for each outbreak, categorized by genus and species of the viruses. The map data of the African continent was employed to draw base maps in the figure. (Available from: https://www.diva-gis.org/).

**Fig 2.**
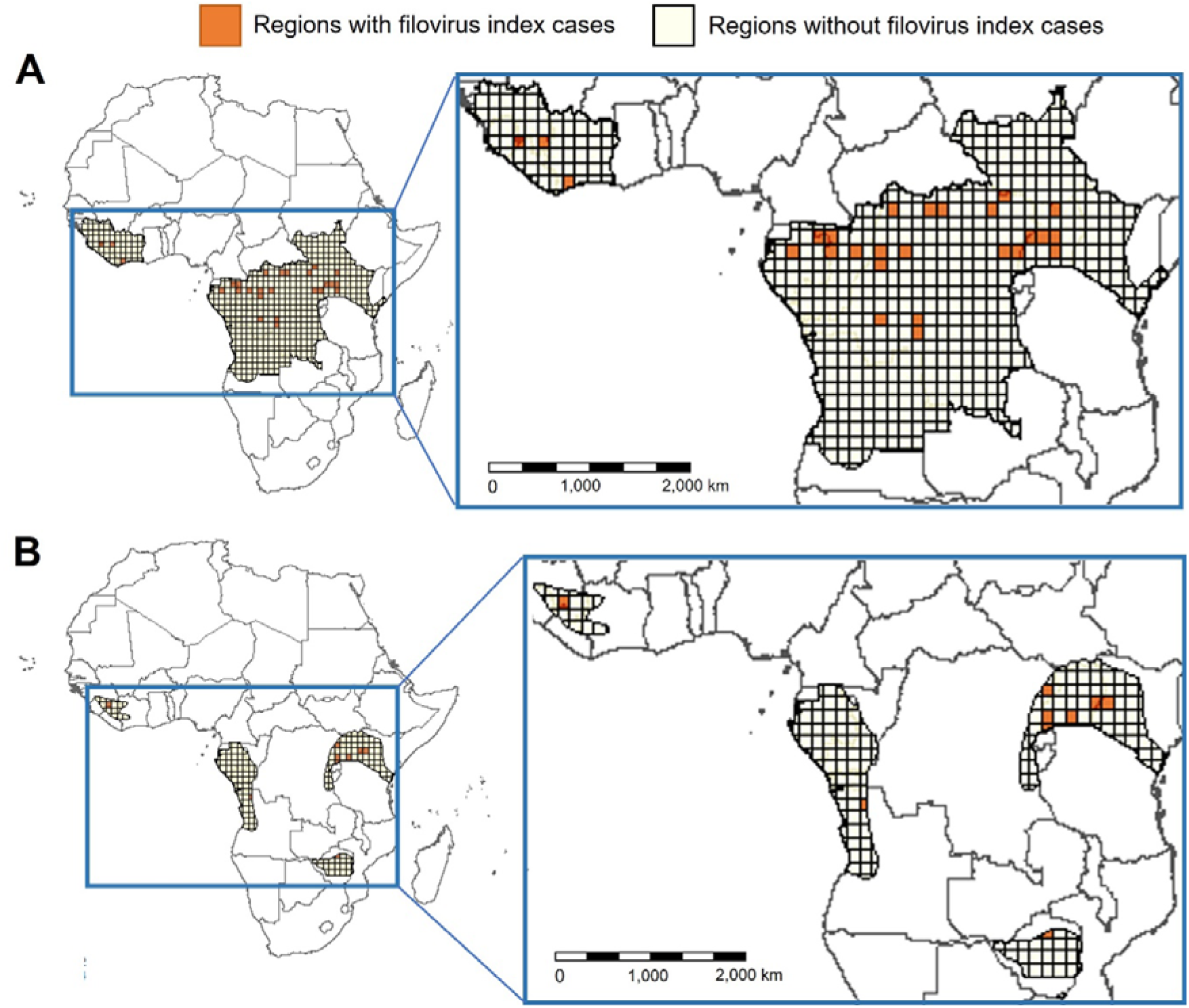
Study area and study units. (A) Shows the study grids with and without *Ebolavirus* index cases. (B) Shows the study grids with and without *Marburgvirus* index cases. The grids with dark orange color represent that the region contains filovirus index cases. The study area is confined to countries with at least one filovirus case. We clipped the area with the reservoir species’ distribution ranges. The map data of the African continent was employed to draw base maps in the figure. (Available from: https://www.diva-gis.org/).

### Descriptive analysis

A total of 524 grids were used for *Ebolavirus* analysis and 197 for *Marburgvirus* analysis. Crude univariate analysis revealed certain characteristics of the study grids (Tables 1, 2). The predator species richness of the orders Accipitriformes and Strigiformes was lower in grids with *Ebolavirus* outbreaks than in those without (Table 1). Also, grids with *Marburgvirus* outbreaks evidenced lower species richness for the predator orders Accipitriformes, Strigiformes, and Carnivora and the family Colubridae (Table 2). All possible predictive variables were included in multivariate logistic models after screening variables with a Pearson correlation coefficient > 0.7 (Supplementary Fig. 1-2). The VIF values were also calculated to assess model multicollinearity. No predictive variable had a VIF > 10.

**Table 1.** Summary of descriptive analysis for *Ebolavirus* index cases.

| Variables | Not occurred<br>(n = 504) | Occurred<br>(n = 20) | Welch's<br>t-test | Wilcox rank<br>sum test |
| --- | --- | --- | --- | --- |
| Avian species richness<br>(order Accipitriformes) | 5.42 ± 7.68 | 3.90 ± 6.11 | 0.29 | 0.27 |
| Avian species richness<br>(order Strigiformes) | 1.46 ± 2.24 | 0.70 ± 1.17 | < 0.05 | < 0.05 |
| Cat species richness<br>(order Carnivora) | 5.01 ± 5.39 | 5.25 ± 7.04 | 0.88 | 0.27 |
| Snake species richness<br>(family colubridae) | 6.73 ± 6.70 | 7.40 ± 8.93 | 0.74 | 0.68 |
| Bat species richness<br>(order Chiroptera) | 10.6 ± 9.17 | 9.65 ± 10.70 | 0.71 | 0.15 |
| Human foot print score | 8.15 ± 4.09 | 9.57 ± 5.48 | 0.26 | 0.41 |
| Elevation | 707.00 ± 402.00 | 701.00 ± 333.00 | 0.93 | 0.99 |
| Precipitation<br>(annual average) | 69.30 ± 47.60 | 116.00 ± 25.20 | <<br>0.001 | < 0.001 |
| Temperature<br>(annual average) | 23.70 ± 2.43 | 23.90 ± 1.39 | 0.69 | 0.88 |
| Population density<br>(per km<sup>2</sup>) | 37.10 ± 66.80 | 65.60 ± 89.70 | 0.18 | 0.06 |
| Gross Domestic Product<br>(per capita) | 2784.00 ±<br>3400.00 | 1733.00 ±<br>2134.00 | < 0.05 | < 0.05 |
| Human Development Index | 0.43 ± 0.07 | 0.41 ± 0.06 | 0.44 | 0.26 |
| Forest cover (%) | 30.70 ± 47.40 | 57.60 ± 45.80 | < 0.05 | < 0.001 |
| Agricultural land use class<br>(% of evergreen broadleaf<br>forests) | 27.50 ± 39.40 | 55.10 ± 43.60 | < 0.05 | < 0.001 |
| Agricultural land use class<br>(% of deciduous broadleaf<br>forests) | 1.07 ± 4.19 | 0.04 ± 0.19 | < 0.05 | 0.06 |
| Agricultural land use class<br>(% of mixed forests) | 0.79 ± 3.50 | 0.19 ± 0.83 | < 0.05 | 0.22 |
| Agricultural land use class<br>(% of woody savannas) | 12.10 ± 21.30 | 13.30 ± 17.50 | 0.78 | 0.46 |
| Agricultural land use class<br>(% of savannas) | 33.70 ± 34.30 | 19.70 ± 26.80 | < 0.05 | < 0.05 |
| Agricultural land use class<br>(% of grasslands) | 20.50 ± 32.30 | 0.60 ± 1.59 | <<br>0.001 | < 0.001 |
| Agricultural land use class<br>(% of permanent wetlands) | 0.35 ± 1.36 | 0.52 ± 1.44 | 0.59 | 0.34 |
| Agricultural land use class<br>(% of croplands) | 1.19 ± 4.71 | 0.37 ± 0.87 | 0.51 | 0.59 |
| Agricultural land use class<br>(% of urban and built-up<br>lands) | $0.07 \pm 0.39$ | $0.20 \pm 0.67$ | 0.41 | 0.14 |
| Agricultural land use class<br>(% of cropland/natural<br>vegetation mosaics) | $1.58 \pm 7.87$ | $9.84 \pm 16.80$ | < 0.05 | < 0.001 |

**Table 2.** Summary of descriptive analysis for *Marburgvirus* index cases.

| Variables | Not occurred<br>(n = 190) | Occurred<br>(n = 7) | Welch's<br>t-test | Wilcox rank<br>sum test |
| --- | --- | --- | --- | --- |
| Bat species richness<br>(order Chiroptera) | 9.09 ± 9.00 | 4.57 ± 3.10 | < 0.05 | 0.11 |
| Avian species richness<br>(order Accipitriformes) | 7.39 ± 10.10 | 1.14 ± 1.86 | < 0.05 | < 0.05 |
| Avian species richness<br>(order Strigiformes) | 2.04 ± 3.05 | 0.14 ± 0.38 | < 0.05 | < 0.05 |
| Cat species richness<br>(order Carnivora) | 4.82 ± 6.90 | 2.57 ± 2.90 | 0.11 | 0.19 |
| Snake species richness<br>(family colubridae) | 5.27 ± 7.15 | 3.00 ± 2.52 | < 0.05 | 0.47 |
| Human foot print score | 10.30 ± 4.47 | 13.90 ± 6.53 | 0.19 | 0.09 |
| Elevation | 778.00 ± 459.00 | 1084.00 ± 453.00 | 0.13 | 0.09 |
| Precipitation (annual<br>average) | 64.70 ± 38.80 | 99.80 ± 32.60 | < 0.05 | < 0.05 |
| Temperature (annual<br>average) | 23.00 ± 2.48 | 22.00 ± 2.02 | 0.24 | 0.21 |
| Population density (per<br>km<sup>2</sup>) | 56.30 ± 83.90 | 157.00 ± 135.00 | < 0.05 | < 0.05 |
| Gross Domestic Product<br>(per capita) | 4085.00 ±<br>4863.00 | 1657.00 ±<br>1536.00 | < 0.05 | < 0.05 |
| Human Development<br>Index | 0.47 ± 0.08 | 0.42 ± 0.04 | < 0.05 | 0.19 |
| Forest cover (%) | 27.10 ± 45.90 | 27.00 ± 37.00 | 0.92 | 0.75 |
| Agricultural land use<br>class 2 (% of evergreen<br>broadleaf forests) | 22.50 ± 34.80 | 24.40 ± 32.40 | 0.82 | 0.67 |
| Agricultural land use<br>class 4 (% of deciduous<br>broadleaf forests) | 0.38 ± 2.68 | 0.00 ± 0.00 | < 0.05 | 0.23 |
| Agricultural land use<br>class 5 (% of mixed<br>forests) | 0.19 ± 1.52 | 0.00 ± 0.00 | 0.09 | 0.41 |
| Agricultural land use<br>class 8 (% of woody<br>savannas) | 8.71 ± 15.70 | 14.40 ± 19.20 | 0.46 | 0.57 |
| Agricultural land use<br>class 9 (% of savannas) | 28.30 ± 31.50 | 28.40 ± 30.10 | 0.89 | 0.58 |
| Agricultural land use<br>class 10 (% of<br>grasslands) | 30.90 ± 32.10 | 4.50 ± 8.41 | <<br>0.001 | < 0.05 |
| Agricultural land use<br>class 11 (% of<br>permanent wetlands) | 0.26 ± 1.42 | 0.65 ± 0.93 | 0.32 | 0.29 |
| Agricultural land use<br>class 12 (% of<br>croplands) | 2.09 ± 6.79 | 3.61 ± 8.43 | 0.65 | 0.23 |
| Agricultural land use<br>class 13 (% of urban and<br>built-up lands) | 0.10 ± 0.42 | 0.46 ± 1.12 | 0.43 | 0.15 |
| Agricultural land use<br>class 14 (% of<br>cropland/natural<br>vegetation mosaics) | 3.99 ± 9.20 | 23.20 ± 24.90 | < 0.05 | < 0.001 |

### Model selection and validation

We constructed saturated models using all available predictive variables and evaluated those variables from an epidemiological perspective (Supplementary Fig. 3). To derive the association between predator species richness and the historical incidence of *Ebolavirus* and *Marburgvirus* in Africa from 2000 to 2021, we used the average values of the predictive variables and the total emergence counts for each grid over that period. At the grid level, the spatial dependencies of filovirus incidences were assessed using the Moran I statistic; the values for *Ebolavirus* and *Marburgvirus* were 0.06 and − 0.03, respectively, when using a row-standardized neighborhood structure (Supplementary Fig. 4-5). The Moran I statistic indicated that only the *Ebolavirus* incidence had a statistically significant spatial or temporal dependency (Supplementary Fig. 4). The Durbin–Watson test results showed that the *Ebolavirus* incidence data were autocorrelated in terms of the residuals of the models, with p-values < 0.05 (Supplementary Table 2). The performances of the models in terms of spatial and spatiotemporal autocorrelations are shown (Supplementary Table 3). Smaller DIC and WAIC values indicate better performance. Based on these results, we fitted Models 1–5 to display the association between predator species richness and the historical incidence of *Ebolavirus*. For *Marburgvirus*, we fitted the model with the average values in line with the Moran I and Durbin– Watson test results.

### Model-estimated association of predator species richness and zoonotic spillover of filoviruses

The results of the final models are shown in Figures 3 and 4. The coefficients and ORs of all covariates are listed (Supplementary Table 4-8). Of all models, Model 2 for *Ebolavirus* exhibited the smallest DIC and WAIC in terms of spatial autocorrelation. In this model, the fourth quartile (OR = 0.04, 95% CI 0.00–0.98) of Strigiformes species richness and the third quartile (OR = 0.15, 95% CI 0.00–0.81) of Colubridae species richness exhibited significantly lower odds of *Ebolavirus* index cases (Figure 3B, Supplementary Table 5). This trend was maintained in Model 5, which showed the lowest DIC and WAIC of all models in terms of spatiotemporal autocorrelation. In this Model, the third quartile (OR = 0.02, 95% CI 0.00–0.84) and fourth quartile (OR = 0.07, 95% CI 0.00–0.42) of Strigiformes species richness, the third quartile (OR = 0.15, 95% CI 0.01–0.73) and fourth quartile (OR = 0.53, 95% CI 0.03–0.85) of Colubridae species richness, and the second quartile (OR = 0.23, 95% CI 0.05–0.94) of Carnivora species richness evidenced significantly lower odds of *Ebolavirus* index cases (Figure 3E, Supplementary Table 8). However, none of the estimated parameters were significant for the other quartiles of Carnivora, Colubridae, and Strigiformes. In the models for *Marburgvirus*, we found no evidence of an association between predator species richness and *Marburgvirus* spillover (Figure 4, Supplementary Table 9). In addition, negative associations between predator species richness and *Ebolavirus* emergence were significant for some of the model quartiles when the “R-INLA” package was used, or when the species richness variables were calculated using the IUCN polygons (Supplementary Fig. 6, Supplementary Table 10-14; Supplementary Fig. 7, Supplementary Table 15-19). Models using the species richness variables including predator species reported to prey on bats did not support any significant association. (Supplementary Fig. 8, Supplementary Table 20-24). No significant association was revealed in the sensitivity analyses for *Marburgvirus* (Supplementary Fig. 9-10, Supplementary Table 25-26).

**Fig 3.**
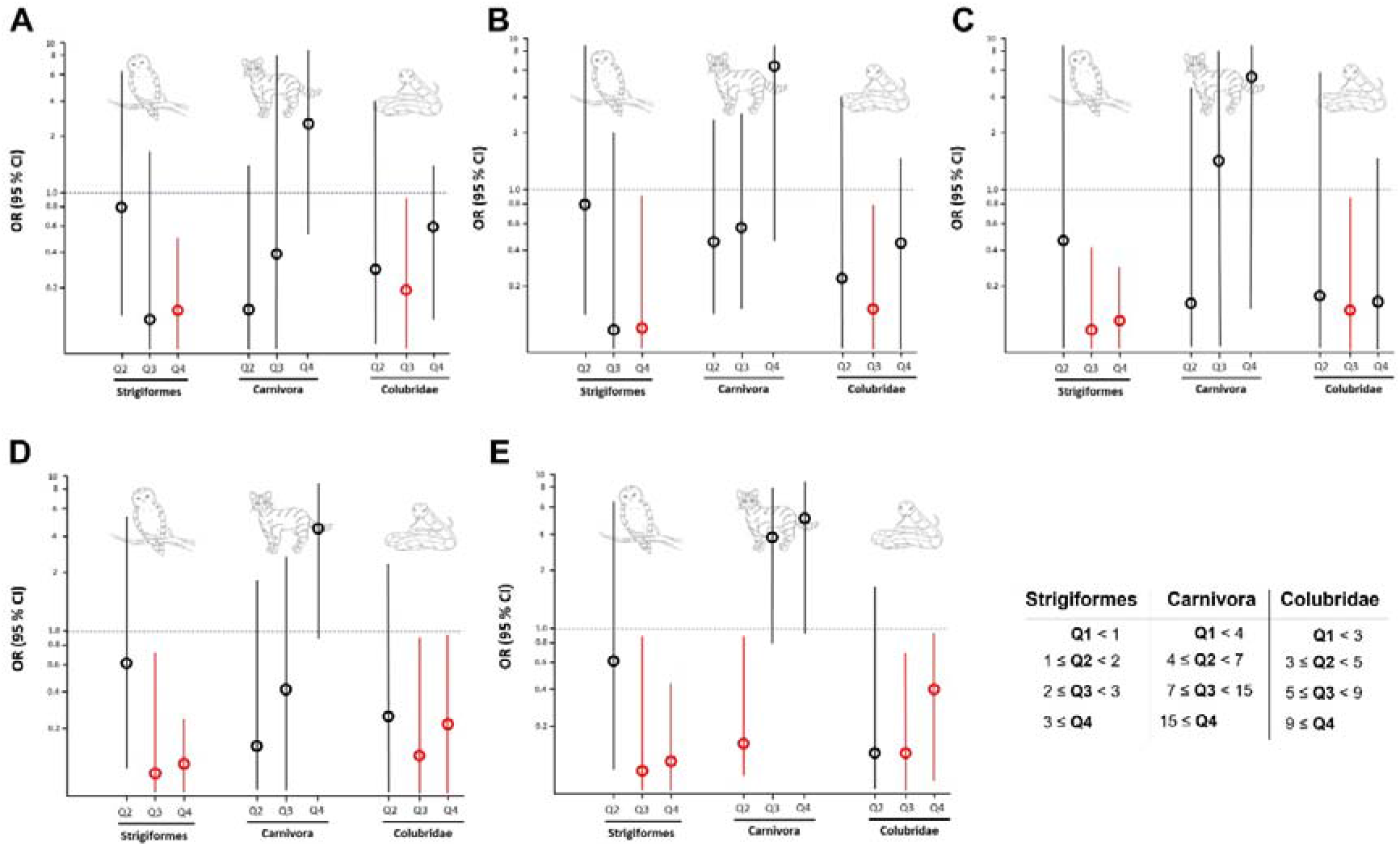
Estimated ORs for *Ebolavirus* incidence according to the degree of species richness. (A) The result of Model 1. (B) The result of Model 2. (C) The result of Model 3. (D) The result of Model 4. (E) The result of Model 5. Model 2 and Model 5 were the best-fitting models with the greatest DIC and WAIC, considering spatial and spatio-temporal autocorrelation, respectively. The dots indicate the estimated ORs, with error bars representing the corresponding 95 % Wald’s credible intervals. Red means that the error bar does not intersect 1. The y-axis is shown on a logarithmic scale. The authors generated draws of each predator.

**Fig 4.**
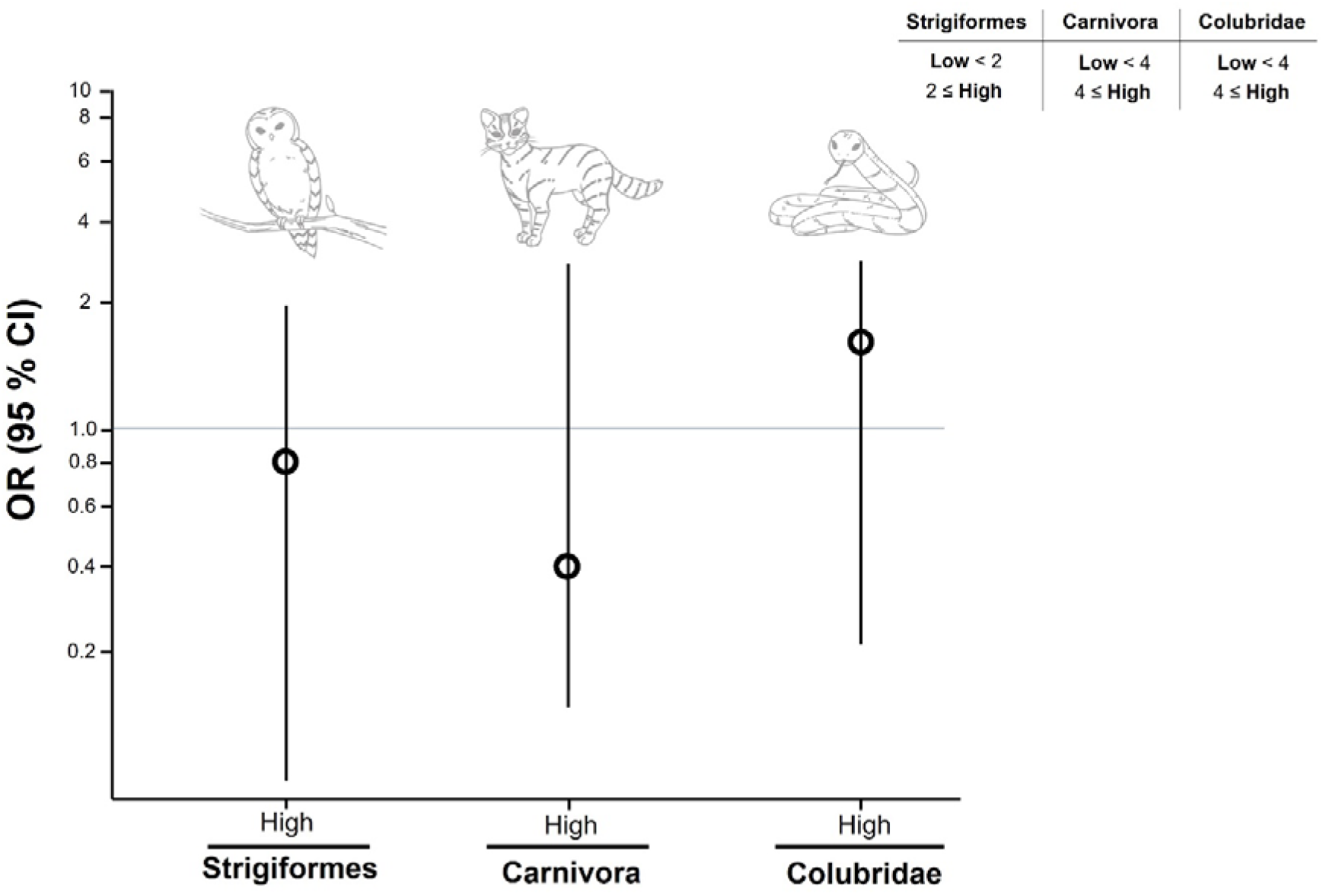
Estimated ORs for *Marburgvirus* incidence according to the degree of species richness. The dots indicate the estimated ORs, with error bars representing the corresponding 95 % Wald’s credible intervals. The y-axis is shown on a logarithmic scale. The authors generated draws of each predator.

## Discussion

We evaluated the association between predator species richness and filovirus spillover in Africa. The results showed that higher species richness in the in the order Strigiformes and family Colubridae was associated with lower odds of *Ebolavirus* spillover compared with that in regions with lower predator species richness. Regardless of the approach taken to calculate species diversity, this association was robust.

The negative association between predator species richness and the risk of *Ebolavirus* spillover suggests top–down regulation of *Ebolavirus* reservoir hosts (i.e., bats) by predators. The crucial roles played by predators in terms of the functional diversity of ecological communities and the control of populations of disease reservoir hosts have been reported previously (9, 10, 12). The greater the predator species richness (i.e., the numbers of predator species within an area), the greater the cascade effect on prey species. The growing body of research on bat predation is slowly improving our understanding of bat predators and the effects of predation on bat populations (12-14). Natural bat predators may include birds, snakes, and mammals. Although few vertebrate predators are known to specialize on bats, and bat predation appears to be mostly opportunistic in nature, generalist and opportunistic predators may substantially impact bat ecology (9, 12, 14) via both direct predation and non-lethal cascade effects, also termed trait-mediated indirect interactions. Thus, predators control the abundance, density, and behavior patterns of prey species, eventually reducing the rate of contact between reservoir hosts and humans and thus mitigating the risk of zoonotic spillover (9). Such suppression is relatively strong in regions wherein ecological diversity is well-maintained.

The predator species richness of the order Strigiformes was significantly and negatively associated with the risk of *Ebolavirus* spillover. This is consistent with previous studies suggesting that owls are primary predators of bats (13, 14). Snakes are also supposed to prey on bats, via two strategies: positioning themselves near bat passage routes (i.e., near the entrances to bat roosts) and entering the refuges (12). Most such behaviors have been reported in tropical regions (29), perhaps because tropical bats roost by hiding among leaves or in open canopies that are accessible to most vertebrate predators. Bat predation is poorly understood; bats fly at night and hide by day. However, it appears that predation of bats by snakes in our study area is more significant than previously thought. More ecological research is required.

Predator species richness was not significantly associated with *Ebolavirus* cases in models that considered only the species reported to prey on bats. This may be attributable to a lack of information on all bat predators. Although the number of known predators is increasing, such research is limited by the ecological characteristics of bats, which render observations of predation difficult (12). Also, *Marburgvirus* occurrences were not consistently associated with predator species richness. The composition of bat species in the *Marburgvirus* regions may explain these results. Given the high bat diversity in the study region, *R. aegypticus*, the primary reservoir host of *Marburgvirus*, would not be the dominant bat species there. Therefore, the extent of predator richness may not have had any discernible effect on bat activities (30). Further studies of bat ecology, diversity, and abundance, especially of *R. aegypticus*, are needed.

Despite the strengths of this ecological study, several limitations should be noted. First, we estimated the diversity of predator species using stacked (aggregated) species distribution models. These models may systematically overestimate site-level species richness (31). Therefore, we adjusted for bias using categorical values of predator species richness. Second, we did not include the temporal variations in species numbers from 2000 to 2021. However, such temporal changes can be ignored because most species considered are classified as IUCN “Least concern” (i.e., low risk of extinction). Third, when measuring species diversity, we simply calculated the numbers of species; we excluded the relative abundances of the predator species. Future research should employ other indicators of diversity such as the Simpson diversity index. Fourth, we considered only three bat species (*E. franqueti, H. monstrosus*, and *M. torquate*) that tested positive by PCR as primary reservoir hosts of *Ebolavirus*. Other probable reservoirs (bat species positive using serological methods) should be included in future studies. Finally, our study units were °× °grids; the use of a different scale (such as 0.5°× 0.5°) could have affected the results. This is the well-known modifiable area unit problem.

The world is still struggling to exit the unprecedented COVID-19 pandemic. It is predicted that the probability of pandemics caused by spillovers may increase in the coming decades, given the tectonic shifts in climate change and anthropogenic environmental degradation. However, although environmental and biodiversity changes may affect the spread of zoonotic diseases via various mechanisms, prevention of outbreaks still depends on containment, i.e., human disease surveillance, vaccines, and therapeutics. Here, we suggest that predator species richness may play a crucial role in mitigating the risk of filovirus spillover. Therefore, attempts to reduce the impacts of zoonotic diseases on public health should incorporate the concept of conservation epidemiology when deriving sustainable solutions that both maintain biodiversity and prevent zoonotic spillover, benefiting both humans and the environment.

## Data Availability

All data produced in the present study are available upon reasonable request to the authors

## Data availability

The datasets used and/or analysed during the current study are available from the corresponding author on reasonable request.

## Code availability

We share the R codes on https://github.com/TaeHChang/R-codes-for-paper-1

## Acknowledgements

This research was supported by a National Research Foundation of Korea (NRF) grant funded by the Korea government (MSIT) (no. NRF-2021R1C1C2012611). The funders had no role in the design and conduct of the study; collection, management, analysis, and interpretation of the data; preparation, review, or approval of the paper; and decision to submit the paper for publication.

## Author contributions

K.D.M. and S.C. conceived, designed, and supervised the study. T.C. collected and analyzed the data.

T.C. wrote the drafts of the paper. K.D.M. and S.C. commented on and revised drafts of the paper. All authors read and approved the final report.

