## supplementary for "Implications of predator species richness in terms of zoonotic spillover transmission of filoviral hemorrhagic fevers in Africa"

**Table of Contents**

**Supplementary Table 1.** Detailed information of covariates used in this study….……………………………… 4

**Supplementary Table 2.** The results of the Durbin-Watson test. ………………………………………………... 4

**Supplementary Table 3.** Assessing the models with DIC and WAIC values. …………………………...…….4-5

**Supplementary Table 4.** Associations of predator species richness with *Ebolavirus* incidence in Model 1. …. …5

**Supplementary Table 5.** Associations of predator species richness with *Ebolavirus* incidence in Model 2..…... 6

**Supplementary Table 6.** Associations of predator species richness with *Ebolavirus* incidence in Model 3.….... 7

**Supplementary Table 7.** Associations of predator species richness with *Ebolavirus* incidence in Model 4…. 7-8

**Supplementary Table 8.** Associations of predator species richness with *Ebolavirus* incidence in Model 5.….8-9

**Supplementary Table 9.** Associations of predator species richness with *Marburgvirus* incidence in the fully adjusted model……………….…………………………………………………………..………………………. .9

**Supplementary Table 10.** Associations of predator species richness with *Ebolavirus* incidence in Model 1. The model parameters were calculated with “R-INLA” package. ………………..…………………………….….…10

**Supplementary Table 11.** Associations of predator species richness with *Ebolavirus* incidence in Model 2. The model parameters were calculated with “R-INLA” package..……………………..……………………….... 10-11

**Supplementary Table 12.** Associations of predator species richness with *Ebolavirus* incidence in Model 3. The model parameters were calculated with “R-INLA” package. ………………………………………...……... 11-12

**Supplementary Table 13.** Associations of predator species richness with *Ebolavirus* incidence in Model 4. The model parameters were calculated with “R-INLA” package.……………………………………..………….12-13

**Supplementary Table 14.** Associations of predator species richness with *Ebolavirus* incidence in Model 5. The model parameters were calculated with “R-INLA” package. ……………………………………………..…….13

**Supplementary Table 15.** Associations of predator species richness with *Ebolavirus* incidence in Model 1, using the species richness variables calculated with IUCN polygons. …………………………………………………14

**Supplementary Table 16.** Associations of predator species richness with *Ebolavirus* incidence in Model 1, using the species richness variables calculated with IUCN polygons. ……………………………………………...14-15

**Supplementary Table 17.** Associations of predator species richness with *Ebolavirus* incidence in Model 3, using the species richness variables calculated with IUCN polygons. …………………………………………..…15-16

**Supplementary Table 18.** Associations of predator species richness with *Ebolavirus* incidence in Model 4, using the species richness variables calculated with IUCN polygons…..………………………………….…….…16-17

**Supplementary Table 19.** Associations of predator species richness with *Ebolavirus* incidence in Model 5, using the species richness variables calculated with IUCN polygons.…………………………………………....……17

**Supplementary Table 20.** Associations of predator species richness with *Ebolavirus* incidence in Model 1, constructed using the species richness variables calculated with Maxent modeling results, and the only species reported to prey on bats were included.……………………….…. ………………………………………………18

**Supplementary Table 21.** Associations of predator species richness with *Ebolavirus* incidence in Model 2, constructed using the species richness variables calculated with Maxent modeling results, and the only species reported to prey on bats were included. . ………………………………….…. ……………………………..18-19

**Supplementary Table 22.** Associations of predator species richness with *Ebolavirus* incidence in Model 3, constructed using the species richness variables calculated with Maxent modeling results, and the only species reported to prey on bats were included..…. ……………………………………………..…..…………………. 19

**Supplementary Table 23.** Associations of predator species richness with *Ebolavirus* incidence in Model 4, constructed using the species richness variables calculated with Maxent modeling results, and the only species reported to prey on bats were included. . ………………………………….…. …………………………………20

**Supplementary Table 24.** Associations of predator species richness with *Ebolavirus* incidence in Model 5, constructed using the species richness variables calculated with Maxent modeling results, and the only species reported to prey on bats were included. ………………………………….…. ………………………………20-21

**Supplementary Table 25.** Associations of predator species richness with *Marburgvirus* incidence in fully adjusted model, using the species richness variables calculated with IUCN polygons. ………………………..…21

**Supplementary Table 26.** Associations of predator species richness with *Marburgvirus* incidence in fully adjusted model, constructed using the species richness variables calculated with Maxent modeling results, and the only species reported to prey on bats were included. …………………………………………………………….22

**Supplementary Figure 1.** Variable collinearity heatmap. Pearson correlation coefficient for a given pair of predictive variables in *Ebolavirus* model. ………………………………………………. ……………………… 23

**Supplementary Figure 2.** Variable collinearity heatmap. Pearson correlation coefficient for a given pair of predictive variables in *Marburgvirus* model. …………………………………………….…………………...… 24

**Supplementary Figure 3.** Directed acyclic graph of the final models in the study……….………. …………….25

**Supplementary Figure 4.** Global Moran’s I plot and Moran’s I statistics for *Ebolavirus* incidence…………… 25

**Supplementary Figure 5.** Global Moran’s I plot and Moran’s I statistics for *Marburgirus* incidence…………. 26

**Supplementary Figure 6.** Estimated ORs for *Ebolavirus* incidence according to the degree of species richness. (A) The result of Model 1. (B) The result of Model 2. (C) The result of Model 3. (D) The result of Model 4. (E) The result of Model 5. The dots indicate the estimated ORs, with error bars representing the corresponding 95 % Wald’s credible intervals. Red means that the error bar does not intersect 1. The models were fitted with “R-INLA” package. The y-axis is shown on a logarithmic scale. The authors generated draws of each predator. …………. 27

**Supplementary Figure 7.** Estimated ORs for *Ebolavirus* incidence according to the degree of species richness. (A) The result of Model 1. (B) The result of Model 2. (C) The result of Model 3. (D) The result of Model 4. (E) The result of Model 5. The dots indicate the estimated ORs, with error bars representing the corresponding 95 % Wald’s credible intervals. Red means that the error bar does not intersect 1. The models were constructed using the species richness variables calculated with IUCN polygons. The y-axis is shown on a logarithmic scale. The authors generated draws of each predator. ………………………………………………………………………. 28

**Supplementary Figure 8.** Estimated ORs for *Ebolavirus* incidence according to the degree of species richness. (A) The result of Model 1. (B) The result of Model 2. (C) The result of Model 3. (D) The result of Model 4. (E) The result of Model 5. The dots indicate the estimated ORs, with error bars representing the corresponding 95 % Wald’s credible intervals. Red means that the error bar does not intersect 1. The reference categories are when the species does not exist. The models were constructed using the species richness variables calculated with Maxent modeling results, and the only species reported to prey on bats were included. The y-axis is shown on a logarithmic scale. The authors generated draws of each predator………………………………………………………..….. 29

**Supplementary Figure 9.** Estimated ORs for *Marburgvirus* incidence according to the degree of species richness. The dots indicate the estimated ORs, with error bars representing the corresponding 95 % Wald’s credible intervals. The model was constructed using the species richness variables calculated with IUCN polygons. The y-axis is shown on a logarithmic scale. The authors generated draws of each predator. ………………………………... 30

**Supplementary Figure 10.** Estimated ORs for *Marburgvirus* incidence according to the degree of species richness. The dots indicate the estimated ORs, with error bars representing the corresponding 95 % Wald’s credible intervals. The reference categories are when the species does not exist. The models were constructed using the species richness variables calculated with Maxent modeling results, and the only species reported to prey on bats were included. The y-axis is shown on a logarithmic scale. The authors generated draws of each predator. …... 31

**Supplementary table 1. Detailed information of covariates used in this study.**

| Variables | Spatial resolution | Temporal range  (resolution) | Calculation method for values in each grid |
| --- | --- | --- | --- |
| Human foot print score | 1 km^2^ | 2000 - 2018 (annual) | average |
| Elevation | 0.008 km^2^ | - | average |
| Precipitation  (annual average) | 20 km^2^ | (monthly average) | average |
| Temperature  (annual average) | 20 km^2^ | (monthly average) | average |
| Population density  (per km^2^) | 1 km^2^ | 2000 - 2020 (annual) | sum |
| Gross Domestic Product (per capita) | 80 km^2^ | 1991 - 2015 (annual) | average |
| Human Development  Index | 80 km^2^ | 1991 - 2015 (annual) | average |
| Forest cover (%) | 0.0009 km^2^ | 2000 - 2021 (annual) | the proportion of forest in each grid |
| Agricultural land use  (% of IGBF class) | 30 km^2^ | 2000 - 2021 (annual) | the proportion of IGBF class in each grid |

**Supplementary table 2. The results of the Durbin-Watson test.**

| **Models** | **lag** | **Autocorrelation** | **D-W statistic** | **p-value** |
| --- | --- | --- | --- | --- |
| *Ebolavirus* model | 1 | 0.13 | 1.73 | < 0.05 |
| *Marburgvirus* model | 1 | -0.03 | 2.06 | 0.87 |

**Supplementary table 3. Assessing the models with DIC and WAIC values.**

| **Model no.** | **Model expression** | **DIC** | **WAIC** |
| --- | --- | --- | --- |
| **1**  **(UH)** | $\ln\left( \frac{p}{1-p} \right) = \beta₀ + \betaₙXₙ + vₙ$ | 225.93 | 251.22 |
| **2**  **(ICAR)** | $\ln\left( \frac{p}{1-p} \right) = \beta₀ + \betaₙXₙ + vₙ + uₙ$ | 224.83 | 240.78 |
| **3**  **(ICAR + time trend)** | $\ln\left( \frac{p}{1-p} \right) = \beta₀ + \betaₙXₙ + vₙ + uₙ + ɑ₁t$ | 309.95 | 330.68 |
| **4**  **(ICAR + randomwalk)** | $\ln\left( \frac{p}{1-p} \right) = \beta₀ + \betaₙXₙ + vₙ + uₙ + ϒₘ$ | 324.37 | 344.70 |
| 5  **(ICAR + random walk +** i**nteraction)** | $\ln\left( \frac{p}{1-p} \right) = \beta₀ + \betaₙXₙ + vₙ + uₙ + ϒₘ + \varphiₙₘ$ | 304.25 | 331.34 |

**Supplementary table 4. Associations of predator species richness with *Ebolavirus* incidence in Model 1.**

| **Variables** | **β (SE)** | **Odds ratio**  **(95 % CI)** |
| --- | --- | --- |
| Species richness of order Strigiformes (no. species) |  |  |
| < 1 | Reference | Reference |
| < 2 | -0.20 | 0.82 (0.07 - 6.16) |
| < 3 | -2.87 | 0.05 (0.00 - 1.67) |
| 3 ≤ | -2.01 | 0.08 (0.00 - 0.49) |
| Species richness of order Carnivora (no. species) |  |  |
| < 4 | Reference | Reference |
| < 7 | -2.32 | 0.09 (0.01 - 1.17) |
| < 15 | -0.88 | 0.41 (0.01 - 8.65) |
| 15 ≤ | 3.76 | 3.19 (0.55 - 9.34) |
| Species richness of family Colubridae (no. species) |  |  |
| < 3 | Reference | Reference |
| < 5 | -1.15 | 0.32 (0.03 - 4.21) |
| < 9 | -1.59 | 0.20 (0.02 - 0.98) |
| 9 ≤ | -0.41 | 0.66 (0.05 - 1.55) |
| Human foot print score | 0.20 | 1.22 (1.09 - 1.44) |
| Precipitation (annual average) | 0.04 | 1.01 (0.99 - 1.04) |
| Temperature (annual average) | -0.30 | 1.02 (1.04 - 1.05) |
| Agricultural land use class (% of deciduous broadleaf forests) | -0.75 | 0.46 (0.39 - 0.57) |
| Agricultural land use class (% of mixed forests) | 0.12 | 1.13 (0.94 - 1.39) |
| Agricultural land use class (% of grasslands) | -0.22 | 0.82 (0.80 - 0.83) |

**Supplementary table 5. Associations of predator species richness with *Ebolavirus* incidence in Model 2.**

| **Variables** | **β (SE)** | **Odds ratio**  **(95 % CI)** |
| --- | --- | --- |
| Species richness of order Strigiformes (no. species) |  |  |
| < 1 | Reference | Reference |
| < 2 | -0.19 | 0.82 (0.04 - 8.85) |
| < 3 | -3.60 | 0.03 (0.00 - 1.97) |
| 3 ≤ | -2.68 | 0.04 (0.00 - 0.98) |
| Species richness of order Carnivora (no. species) |  |  |
| < 4 | Reference | Reference |
| < 7 | -2.21 | 0.56 (0.07 - 3.06) |
| < 15 | -0.58 | 0.63 (0.10 - 2.71) |
| 15 ≤ | 1.98 | 7.21 (0.44 - 9.55) |
| Species richness of family Colubridae (no. species) |  |  |
| < 3 | Reference | Reference |
| < 5 | -1.44 | 0.23 (0.01 - 5.38) |
| < 9 | -1.91 | 0.15 (0.00 - 0.81) |
| 9 ≤ | -0.77 | 0.46 (0.01 - 1.09) |
| Human foot print score | 0.22 | 1.25 (0.90 - 1.85) |
| Precipitation (annual average) | 0.02 | 1.02 (0.98 - 1.03) |
| Temperature (annual average) | 0.03 | 1.03 (0.45 - 2.51) |
| Agricultural land use class (% of deciduous broadleaf forests) | -2.93 | 0.05 (0.00 - 0.48) |
| Agricultural land use class (% of mixed forests) | -0.20 | 0.82 (0.25 - 1.61) |
| Agricultural land use class (% of grasslands) | -0.47 | 0.62 (0.38 - 0.87) |

**Supplementary table 6. Associations of predator species richness with *Ebolavirus* incidence in Model 3.**

| **Variables** | **β (SE)** | | **Odds ratio**  **(95 % CI)** |
| --- | --- | --- | --- |
| Species richness of order Strigiformes (no. species) |  | |  |
| < 1 | Reference | | Reference |
| < 2 | -0.78 | | 0.45 (0.02 - 9.94) |
| < 3 | -3.86 | | 0.04 (0.00 - 0.43) |
| 3 ≤ | -2.19 | | 0.09 (0.00 - 0.33) |
| Species richness of order Carnivora (no. species) |  | |  |
| < 4 | Reference | | Reference |
| < 7 | -1.89 | | 0.15 (0.02 - 5.95) |
| < 15 | 0.52 | | 1.69 (0.02 - 8.53) |
| 15 ≤ | 1.87 | | 6.48 (0.87 - 9.52) |
| Species richness of family Colubridae (no. species) |  | |  |
| < 3 | Reference | | Reference |
| < 5 | -1.75 | | 0.17 (0.01 - 6.68) |
| < 9 | -1.99 | | 0.13 (0.00 - 0.95) |
| 9 ≤ | -1.81 | | 0.16 (0.00 - 1.21) |
| Human foot print score | 0.29 | | 1.33 (1.07 - 1.61) |
| Precipitation (annual average) | 0.06 | | 1.06 (1.03 - 1.09) |
| Temperature (annual average) | 0.07 | | 1.07 (0.50 - 2.30) |
| Agricultural land use class (% of deciduous broadleaf forests) | -0.43 | | 0.64 (0.33 - 1.16) |
| Agricultural land use class (% of mixed forests) | 0.01 | | 1.00 (0.65 - 1.26) |
| Agricultural land use class (% of grasslands) | -0.26 | 0.77 (0.58 - 0.98) | |

**Supplementary table 7. Associations of predator species richness with *Ebolavirus* incidence in Model 4.**

| **Variables** | **β (SE)** | | | **Odds ratio**  **(95 % CI)** |
| --- | --- | --- | --- | --- |
| Species richness of order Strigiformes (no. species) |  | | |  |
| < 1 | Reference | | | Reference |
| < 2 | -0.51 | | | 0.65 (0.12 - 5.72) |
| < 3 | -3.04 | | | 0.05 (0.00 - 0.79) |
| 3 ≤ | -2.75 | | | 0.08 (0.00 - 0.23) |
| Species richness of order Carnivora (no. species) |  | | |  |
| < 4 | Reference | | | Reference |
| < 7 | -1.76 | | | 0.17 (0.01 - 1.58) |
| < 15 | -0.83 | | | 0.43 (0.03 - 3.41) |
| 15 ≤ | 1.69 | | | 5.46 (0.97 - 8.23) |
| Species richness of family Colubridae (no. species) |  | | |  |
| < 3 | Reference | | | Reference |
| < 5 | -1.16 | | | 0.31 (0.03 - 3.30) |
| < 9 | -2.22 | | | 0.11 (0.01 - 0.89) |
| 9 ≤ | -1.38 | | | 0.25 (0.02 - 0.94) |
| Human foot print score | 0.20 | | | 1.23 (1.09 - 1.35) |
| Precipitation (annual average) | 0.03 | | | 1.02 (0.98 - 1.03) |
| Temperature (annual average) | -0.15 | | | 0.88 (0.60 - 1.30) |
| Agricultural land use class (% of deciduous broadleaf forests) | -1.14 | | | 0.64 (0.05 - 2.77) |
| Agricultural land use class (% of mixed forests) | -0.07 | | | 1.01 (0.01 - 1.26) |
| Agricultural land use class (% of grasslands) | | -0.21 | 0.80 (0.56 - 0.99) | |

**Supplementary table 8. Associations of predator species richness with *Ebolavirus* incidence in Model 5.**

| **Variables** | **β (SE)** | | | **Odds ratio**  **(95 % CI)** |
| --- | --- | --- | --- | --- |
| Species richness of order Strigiformes (no. species) |  | | |  |
| < 1 | Reference | | | Reference |
| < 2 | -0.26 | | | 0.77 (0.05 - 6.58) |
| < 3 | -3.51 | | | 0.02 (0.00 - 0.84) |
| 3 ≤ | -2.70 | | | 0.07 (0.00 - 0.42) |
| Species richness of order Carnivora (no. species) |  | | |  |
| < 4 | Reference | | | Reference |
| < 7 | -1.45 | | | 0.23 (0.05 - 0.94) |
| < 15 | 1.37 | | | 3.93 (0.78 - 7.97) |
| 15 ≤ | 1.92 | | | 6.82 (0.98 - 9.55) |
| Species richness of family Colubridae (no. species) |  | | |  |
| < 3 | Reference | | | Reference |
| < 5 | -2.03 | | | 0.13 (0.00 - 1.46) |
| < 9 | -1.87 | | | 0.15 (0.01 - 0.73) |
| 9 ≤ | -0.62 | | | 0.53 (0.03 - 0.84) |
| Human foot print score | 0.33 | | | 1.39 (0.86 - 1.99) |
| Precipitation (annual average) | 0.12 | | | 1.13 (1.07 - 1.21) |
| Temperature (annual average) | -0.17 | | | 0.84 (0.37 - 2.64) |
| Agricultural land use class (% of deciduous broadleaf forests) | -0.01 | | | 0.99 (0.48 - 1.78) |
| Agricultural land use class (% of mixed forests) | -0.09 | | | 0.91 (0.28 - 2.61) |
| Agricultural land use class (% of grasslands) | | -0.33 | 0.72 (0.60 - 0.85) | |

**Supplementary table 9. Associations of predator species richness with *Marburgvirus* incidence in the fully adjusted model.**

| **Variables** | **β (SE)** | **Odds ratio (95 % CI)** |
| --- | --- | --- |
| Species richness of order Strigiformes (no. species) |  |  |
| < 2 | Reference | Reference |
| 2 ≤ | -0.24 (0.34) | 0.81 (0.01 - 1.97) |
| Species richness of order Carnivora (no. species) |  |  |
| < 3 | Reference | Reference |
| 3 ≤ | -0.83 (1.62) | 0.43 (0.10 - 2.55) |
| Species richness of family Colubridae (no. species) |  |  |
| < 4 | Reference | Reference |
| 4 ≤ | 0.57 (0.78) | 1.67 (0.20 - 2.75) |
| Human foot print score | -0.05 (0.16) | 1.09 (0.92 - 1.29) |
| Precipitation (annual average) | 0.01 (0.01) | 1.02 (0.98 - 1.03) |
| Temperature (annual average) | -0.32 (0.23) | 0.88 (0.60 - 1.30) |
| Gross Domestic Product (per capita) | 0.00 (0.00) | 1.00 (1.00 -1.00) |
| Population density (per km^2) | 0.01 (0.01) | 1.00 (0.98 - 1.01) |
| Agricultural land use class (% of deciduous broadleaf forests) | -0.30 (5.62) | 0.54 (0.01 - 1.07) |
| Agricultural land use class (% of mixed forests) | -0.56 (4.14) | 1.03 (0.01 - 1.26) |
| Agricultural land use class (% of savannas) | -0.02 (0.02) | 0.99 (0.96 - 1.02) |
| Agricultural land use class (% of grasslands) | -0.06 (0.05) | 0.82 (0.39 - 0.98) |
| Agricultural land use class (% of cropland/natural vegetation mosaics) | 0.03 (0.03) | 1.01 (0.96 - 1.10) |

**Supplementary table 10. Associations of predator species richness with *Ebolavirus* incidence in Model 1. The model parameters were calculated with “R-INLA” package.**

| **Variables** | **β (SE)** | | **Odds ratio**  **(95 % CI)** |
| --- | --- | --- | --- |
| Species richness of order Strigiformes (no. species) |  | |  |
| < 1 | Reference | | Reference |
| < 2 | 0.56 (0.62) | | 1.81 (0.46 - 5.51) |
| < 3 | -1.89 (1.64) | | 0.18 (0.01 - 2.41) |
| 3 ≤ | -2.71 (1.80) | | 0.08 (0.01 - 1.26) |
| Species richness of order Carnivora (no. species) |  | |  |
| < 4 | Reference | | Reference |
| < 7 | -1.33 (0.72) | | 0.28 (0.05 - 1.06) |
| < 15 | -0.48 (0.89) | | 0.64 (0.09 - 3.21) |
| 15 ≤ | 2.32 (1.61) | | 7.03 (0.61 - 9.96) |
| Species richness of family Colubridae (no. species) |  | |  |
| < 3 | Reference | | Reference |
| < 5 | -0.97 (0.72) | | 0.39 (0.08 - 1.44) |
| < 9 | -1.66 (0.70) | | 0.19 (0.05 - 0.76) |
| 9 ≤ | -0.76 (0.78) | | 0.47 (0.09 - 2.27) |
| Human foot print score | 0.13 (0.10) | | 1.14 (0.95 - 1.39) |
| Precipitation (annual average) | 0.01 (0.01) | | 1.01 (0.99 - 1.04) |
| Temperature (annual average) | -0.04 (0.20) | | 0.95 (0.65 - 1.45) |
| Agricultural land use class (% of deciduous broadleaf forests) | -1.06 (1.14) | | 0.40 (0.03 - 2.16) |
| Agricultural land use class (% of mixed forests) | 0.04 (0.22) | | 1.07 (0.63 - 1.48) |
| Agricultural land use class (% of grasslands) | -0.38 (0.18) | 0.69 (0.46 - 0.94) | |

**Supplementary table 11. Associations of predator species richness with *Ebolavirus* incidence in Model 2. The model parameters were calculated with “R-INLA” package.**

| **Variables** | **β (SE)** | | **Odds ratio**  **(95 % CI)** |
| --- | --- | --- | --- |
| Species richness of order Strigiformes (no. species) |  | |  |
| < 1 | Reference | | Reference |
| < 2 | 0.69 (0.53) | | 2.01 (0.70 - 5.69) |
| < 3 | -1.75 (1.50) | | 0.19 (0.01 - 2.21) |
| 3 ≤ | -2.39 (1.56) | | 0.10 (0.01 - 1.33) |
| Species richness of order Carnivora (no. species) |  | |  |
| < 4 | Reference | | Reference |
| < 7 | -1.29 (0.77) | | 0.29 (0.05 - 0.96) |
| < 15 | -0.50 (0.83) | | 0.63 (0.10 - 2.71) |
| 15 ≤ | 2.03 (1.34) | | 7.26 (0.58 - 13.55) |
| Species richness of family Colubridae (no. species) |  | |  |
| < 3 | Reference | | Reference |
| < 5 | -0.95 (0.65) | | 0.40 (0.09 - 1.38) |
| < 9 | -1.71 (0.65) | | 0.18 (0.05 - 0.61) |
| 9 ≤ | -0.81 (0.62) | | 0.45 (0.10 - 1.07) |
| Human foot print score | 0.20 (0.05) | | 1.23 (1.09 - 1.35) |
| Precipitation (annual average) | 0.01 (0.01) | | 1.02 (0.98 - 1.03) |
| Temperature (annual average) | -0.04 (0.16) | | 0.88 (0.60 - 1.30) |
| Agricultural land use class (% of deciduous broadleaf forests) | -0.60 (0.98) | | 0.64 (0.05 - 2.77) |
| Agricultural land use class (% of mixed forests) | 0.05 (0.17) | | 1.03 (0.01 - 1.26) |
| Agricultural land use class (% of grasslands) | -0.24 (0.15) | 0.80 (0.56 - 0.99) | |

**Supplementary table 12. Associations of predator species richness with *Ebolavirus* incidence in Model 3. The model parameters were calculated with “R-INLA” package.**

| **Variables** | **β (SE)** | | **Odds ratio**  **(95 % CI)** |
| --- | --- | --- | --- |
| Species richness of order Strigiformes (no. species) |  | |  |
| < 1 | Reference | | Reference |
| < 2 | 0.64 (0.49) | | 1.89 (0.71 - 4.94) |
| < 3 | -1.54 (1.29) | | 0.23 (0.01 - 2.03) |
| 3 ≤ | -1.02 (1.56) | | 0.39 (0.04 - 2.33) |
| Species richness of order Carnivora (no. species) |  | |  |
| < 4 | Reference | | Reference |
| < 7 | -1.38 (0.77) | | 0.26 (0.05 - 0.95) |
| < 15 | -0.09 (0.77) | | 0.94 (0.19 - 3.66) |
| 15 ≤ | 1.78 (0.94) | | 6.06 (0.87 - 8.52) |
| Species richness of family Colubridae (no. species) |  | |  |
| < 3 | Reference | | Reference |
| < 5 | -0.54 (0.55) | | 0.59 (0.18 - 1.68) |
| < 9 | -1.59 (0.57) | | 0.21 (0.06 - 0.61) |
| 9 ≤ | -0.82 (0.62) | | 0.44 (0.12 - 1.01) |
| Human foot print score | 0.19 (0.05) | | 1.23 (1.07 - 1.32) |
| Precipitation (annual average) | 0.01 (0.01) | | 1.02 (0.99 - 1.03) |
| Temperature (annual average) | -0.04 (0.16) | | 0.88 (0.60 - 1.30) |
| Agricultural land use class (% of deciduous broadleaf forests) | -0.25 (0.45) | | 0.83 (0.27 - 1.55) |
| Agricultural land use class (% of mixed forests) | -0.02 (0.18) | | 1.00 (0.65 - 1.26) |
| Agricultural land use class (% of grasslands) | -0.03 (0.02) | 0.74 (0.55 - 0.92) | |

**Supplementary table 13. Associations of predator species richness with *Ebolavirus* incidence in Model 4. The model parameters were calculated with “R-INLA” package.**

| **Variables** | **β (SE)** | | **Odds ratio**  **(95 % CI)** |
| --- | --- | --- | --- |
| Species richness of order Strigiformes (no. species) |  | |  |
| < 1 | Reference | | Reference |
| < 2 | 0.64 (0.49) | | 1.90 (0.72 - 4.95) |
| < 3 | -1.55 (1.29) | | 0.23 (0.01 - 2.04) |
| 3 ≤ | -1.04 (1.05) | | 0.38 (0.03 - 2.33) |
| Species richness of order Carnivora (no. species) |  | |  |
| < 4 | Reference | | Reference |
| < 7 | -1.38 (0.77) | | 0.27 (0.04 - 0.95) |
| < 15 | -0.11 (0.75) | | 0.92 (0.18 - 3.59) |
| 15 ≤ | 1.78 (0.95) | | 6.10 (0.87 - 8.55) |
| Species richness of family Colubridae (no. species) |  | |  |
| < 3 | Reference | | Reference |
| < 5 | -0.55 (0.56) | | 0.59 (0.18 - 1.66) |
| < 9 | -1.59 (0.58) | | 0.21 (0.06 - 0.61) |
| 9 ≤ | -0.81 (0.52) | | 0.44 (0.12 - 0.91) |
| Human foot print score | 0.19 (0.05) | | 1.23 (1.09 - 1.35) |
| Precipitation (annual average) | 0.01 (0.01) | | 1.02 (0.98 - 1.03) |
| Temperature (annual average) | -0.04 (0.16) | | 0.88 (0.60 - 1.30) |
| Agricultural land use class (% of deciduous broadleaf forests) | -0.26 (0.45) | | 0.64 (0.05 - 2.77) |
| Agricultural land use class (% of mixed forests) | -0.02 (0.17) | | 1.01 (0.01 - 1.26) |
| Agricultural land use class (% of grasslands) | -0.31 (0.15) | 0.80 (0.56 - 0.99) | |

**Supplementary table 14. Associations of predator species richness with *Ebolavirus* incidence in Model 5. The model parameters were calculated with “R-INLA” package.**

| **Variables** | **β (SE)** | | **Odds ratio**  **(95 % CI)** |
| --- | --- | --- | --- |
| Species richness of order Strigiformes (no. species) |  | |  |
| < 1 | Reference | | Reference |
| < 2 | 0.64 (0.49) | | 1.89 (0.72 - 4.96) |
| < 3 | -1.55 (1.29) | | 0.23 (0.01 - 2.04) |
| 3 ≤ | -1.04 (1.05) | | 0.38 (0.04 - 2.33) |
| Species richness of order Carnivora (no. species) |  | |  |
| < 4 | Reference | | Reference |
| < 7 | -1.39 (0.77) | | 0.26 (0.05 - 0.94) |
| < 15 | -0.11 (0.75) | | 0.92 (0.18 - 3.97) |
| 15 ≤ | 1.79 (0.98) | | 6.10 (0.87 - 36.55) |
| Species richness of family Colubridae (no. species) |  | |  |
| < 3 | Reference | | Reference |
| < 5 | -0.55 (0.56) | | 0.59 (0.18 - 1.68) |
| < 9 | -1.58 (0.57) | | 0.21 (0.06 - 0.61) |
| 9 ≤ | -0.82 (0.64) | | 0.44 (0.12 - 0.75) |
| Human foot print score | 0.20 (0.05) | | 1.23 (1.09 - 1.35) |
| Precipitation (annual average) | 0.01 (0.01) | | 1.02 (0.98 - 1.03) |
| Temperature (annual average) | -0.12 (0.13) | | 0.88 (0.60 - 1.30) |
| Agricultural land use class (% of deciduous broadleaf forests) | -0.20 (0.39) | | 0.84 (0.33 - 1.49) |
| Agricultural land use class (% of mixed forests) | 0.01 (0.14) | | 1.03 (0.72 - 1.26) |
| Agricultural land use class (% of grasslands) | -0.24 (0.15) | 0.60 (0.60 - 0.97) | |

**Supplementary table 15. Associations of predator species richness with *Ebolavirus* incidence in Model 1, using the species richness variables calculated with IUCN polygons.**

| **Variables** | **β (SE)** | | **Odds ratio**  **(95 % CI)** |
| --- | --- | --- | --- |
| Species richness of order Strigiformes (no. species) |  | |  |
| < 6 | Reference | | Reference |
| < 8 | 0.28 | | 1.33 (0.40 - 3.26) |
| < 9 | -0.35 | | 0.70 (0.15 - 5.23) |
| 9 ≤ | 0.31 | | 1.36 (0.73 - 2.17) |
| Species richness of order Carnivora (no. species) |  | |  |
| < 13 | Reference | | Reference |
| < 17 | -0.37 | | 0.69 (0.09 - 6.25) |
| < 19 | -0.41 | | 0.66 (0.01 - 9.04) |
| 19 ≤ | 1.10 | | 2.79 (0.56 - 6.78) |
| Species richness of family Colubridae (no. species) |  | |  |
| < 20 | Reference | | Reference |
| < 23 | -0.23 | | 0.79 (0.61 - 0.98) |
| 23 ≤ | -0.26 | | 0.77 (0.62 - 1.05) |
| Human foot print score | 0.02 | | 1.02 (0.86 - 1.23) |
| Precipitation (annual average) | 0.03 | | 1.02 (0.99 - 1.07) |
| Temperature (annual average) | 0.28 | | 1.33 (0.80 - 2.41) |
| Agricultural land use class (% of deciduous broadleaf forests) | -1.37 | | 0.25 (0.02 - 2.90) |
| Agricultural land use class (% of mixed forests) | -0.07 | | 0.93 (0.36 - 1.63) |
| Agricultural land use class (% of grasslands) | -0.33 | 0.71 (0.55 - 0.94) | |

**Supplementary table 16. Associations of predator species richness with *Ebolavirus* incidence in Model 2, using the species richness variables calculated with IUCN polygons.**

| **Variables** | **β (SE)** | | **Odds ratio**  **(95 % CI)** |
| --- | --- | --- | --- |
| Species richness of order Strigiformes (no. species) |  | |  |
| < 6 | Reference | | Reference |
| < 8 | 0.27 | | 1.30 (0.86 - 3.57) |
| < 9 | -0.30 | | 0.72 (0.35 - 2.23) |
| 9 ≤ | -0.29 | | 0.73 (0.13 - 0.97) |
| Species richness of order Carnivora (no. species) |  | |  |
| < 13 | Reference | | Reference |
| < 17 | 0.31 | | 1.35 (0.04 - 2.01) |
| < 19 | -0.49 | | 0.61 (0.01 - 4.94) |
| 19 ≤ | 1.42 | | 4.13 (0.88 - 9.78) |
| Species richness of family Colubridae (no. species) |  | |  |
| < 20 | Reference | | Reference |
| < 23 | -0.28 | | 0.76 (0.68 - 0.99) |
| 23 ≤ | -0.35 | | 0.66 (0.22 - 0.93) |
| Human foot print score | 0.04 | | 1.04 (0.88 - 1.28) |
| Precipitation (annual average) | 0.03 | | 1.03 (1.00 - 1.06) |
| Temperature (annual average) | 0.05 | | 1.05 (0.57 - 1.99) |
| Agricultural land use class (% of deciduous broadleaf forests) | -0.82 | | 0.43 (0.12 - 1.07) |
| Agricultural land use class (% of mixed forests) | -0.08 | | 0.92 (0.29 - 1.54) |
| Agricultural land use class (% of grasslands) | -0.91 | 0.40 (0.31 - 0.56) | |

**Supplementary table 17. Associations of predator species richness with *Ebolavirus* incidence in Model 3, using the species richness variables calculated with IUCN polygons.**

| **Variables** | **β (SE)** | | **Odds ratio**  **(95 % CI)** |
| --- | --- | --- | --- |
| Species richness of order Strigiformes (no. species) |  | |  |
| < 6 | Reference | | Reference |
| < 8 | 0.21 | | 1.30 (0.86 - 4.26) |
| < 9 | -0.28 | | 0.75 (0.55 - 1.09) |
| 9 ≤ | -0.22 | | 0.80 (0.53 - 0.97) |
| Species richness of order Carnivora (no. species) |  | |  |
| < 13 | Reference | | Reference |
| < 17 | 0.86 | | 2.38 (0.54 - 9.01) |
| < 19 | -0.84 | | 0.43 (0.01 - 5.74) |
| 19 ≤ | 1.63 | | 5.10 (0.98 - 9.78) |
| Species richness of family Colubridae (no. species) |  | |  |
| < 20 | Reference | | Reference |
| < 23 | -0.17 | | 0.84 (0.61 - 0.98) |
| 23 ≤ | -0.18 | | 0.84 (0.62 - 1.03) |
| Human foot print score | -0.02 | | 0.98 (0.88 - 1.08) |
| Precipitation (annual average) | 0.03 | | 1.03 (1.01 - 1.06) |
| Temperature (annual average) | 0.11 | | 1.10 (0.75 - 1.63) |
| Agricultural land use class (% of deciduous broadleaf forests) | -0.32 | | 0.72 (0.35 - 1.14) |
| Agricultural land use class (% of mixed forests) | -0.03 | | 0.97 (0.55 - 1.28) |
| Agricultural land use class (% of grasslands) | -0.21 | 0.81 (0.67 - 0.92) | |

**Supplementary table 18. Associations of predator species richness with *Ebolavirus* incidence in Model 4, using the species richness variables calculated with IUCN polygons.**

| **Variables** | **β (SE)** | | **Odds ratio**  **(95 % CI)** |
| --- | --- | --- | --- |
| Species richness of order Strigiformes (no. species) |  | |  |
| < 6 | Reference | | Reference |
| < 8 | 0.49 | | 1.63 (0.86 - 4.06) |
| < 9 | 0.18 | | 1.01 (0.85 - 2.23) |
| 9 ≤ | -0.50 | | 0.60 (0.33 - 0.97) |
| Species richness of order Carnivora (no. species) |  | |  |
| < 13 | Reference | | Reference |
| < 17 | 0.76 | | 2.14 (0.41 - 7.58) |
| < 19 | -3.23 | | 0.04 (0.00 - 0.56) |
| 19 ≤ | 0.69 | | 1.99 (0.84 - 5.78) |
| Species richness of family Colubridae (no. species) |  | |  |
| < 20 | Reference | | Reference |
| < 23 | -0.50 | | 0.61 (0.21 - 0.95) |
| 23 ≤ | -0.51 | | 0.60 (0.18 - 0.98) |
| Human foot print score | -0.09 | | 0.92 (0.72 - 1.21) |
| Precipitation (annual average) | 0.05 | | 1.05 (1.01 - 1.09) |
| Temperature (annual average) | 0.23 | | 1.26 (0.48 - 4.33) |
| Agricultural land use class (% of deciduous broadleaf forests) | -0.30 | | 0.74 (0.39 - 1.38) |
| Agricultural land use class (% of mixed forests) | 0.17 | | 1.19 (0.72 - 1.77) |
| Agricultural land use class (% of grasslands) | -0.74 | 0.47 (0.40 - 0.57) | |

**Supplementary table 19. Associations of predator species richness with *Ebolavirus* incidence in Model 5, using the species richness variables calculated with IUCN polygons.**

| **Variables** | **β (SE)** | | **Odds ratio**  **(95 % CI)** |
| --- | --- | --- | --- |
| Species richness of order Strigiformes (no. species) |  | |  |
| < 6 | Reference | | Reference |
| < 8 | 0.38 | | 1.46 (0.91 - 3.96) |
| < 9 | -0.44 | | 0.64 (0.55 - 0.94) |
| 9 ≤ | -0.37 | | 0.68 (0.51 - 0.96) |
| Species richness of order Carnivora (no. species) |  | |  |
| < 13 | Reference | | Reference |
| < 17 | 0.94 | | 2.55 (0.94 - 7.01) |
| < 19 | -0.21 | | 0.81 (0.61 - 2.02) |
| 19 ≤ | 0.62 | | 1.86 (0.84 - 2.78) |
| Species richness of family Colubridae (no. species) |  | |  |
| < 20 | Reference | | Reference |
| < 23 | -0.37 | | 0.69 (0.41 - 1.02) |
| 23 ≤ | -0.40 | | 0.66 (0.32 - 0.99) |
| Human foot print score | -0.06 | | 0.94 (0.80 - 1.10) |
| Precipitation (annual average) | 0.04 | | 1.04 (1.02 - 1.07) |
| Temperature (annual average) | 0.17 | | 1.19 (0.67 - 1.97) |
| Agricultural land use class (% of deciduous broadleaf forests) | -0.27 | | 0.77 (0.42 - 1.26) |
| Agricultural land use class (% of mixed forests) | -0.08 | | 0.92 (0.39 - 1.41) |
| Agricultural land use class (% of grasslands) | -0.56 | 0.57 (0.52 - 0.65) | |

**Supplementary table 20. Associations of predator species richness with *Ebolavirus* incidence in Model 1, constructed using the species richness variables calculated with Maxent modeling results, and the only species reported to prey on bats were included.**

| **Variables** | **β (SE)** | **Odds ratio**  **(95 % CI)** |
| --- | --- | --- |
| Presence of order Strigiformes |  |  |
| without the predator | Reference | Reference |
| with the predator | -0.20 | 0.82 (0.19 - 2.86) |
| Presence of order Carnivora |  |  |
| without the predator | Reference | Reference |
| with the predator | -2.14 | 0.13 (0.01 - 1.29) |
| Presence of order Squamata |  |  |
| without the predator | Reference | Reference |
| with the predator | 1.46 | 4.32 (0.35 - 5.84) |
| Bat species richness (order Chiroptera) | -0.02 | 0.97 (0.93 - 1.15) |
| Human foot print score | -0.02 | 0.97 (0.91 - 1.12) |
| Precipitation (annual average) | 0.01 | 1.02 (0.98 - 1.03) |
| Temperature (annual average) | -0.16 | 0.88 (0.60 - 1.30) |
| Agricultural land use class (% of deciduous broadleaf forests) | -0.61 | 0.54 (0.01 - 1.07) |
| Agricultural land use class (% of mixed forests) | 0.03 | 1.03 (0.01 - 1.26) |
| Agricultural land use class (% of grasslands) | -0.20 | 0.82 (0.39 - 0.98) |

**Supplementary table 21. Associations of predator species richness with *Ebolavirus* incidence in Model 2, constructed using the species richness variables calculated with Maxent modeling results, and the only species reported to prey on bats were included.**

| **Variables** | **β (SE)** | **Odds ratio**  **(95 % CI)** |
| --- | --- | --- |
| Presence of order Strigiformes |  |  |
| without the predator | Reference | Reference |
| with the predator | 0.46 | 1.61 (0.62 - 3.84) |
| Presence of order Carnivora |  |  |
| without the predator | Reference | Reference |
| with the predator | -1.86 | 0.17 (0.01 - 1.42) |
| Presence of order Squamata |  |  |
| without the predator | Reference | Reference |
| with the predator | 1.63 | 5.73 (0.57 - 9.59) |
| Bat species richness (order Chiroptera) | -0.05 | 0.95 (0.86 - 1.03) |
| Human foot print score | -0.01 | 0.98 (0.91 - 1.12) |
| Precipitation (annual average) | 0.01 | 1.02 (0.98 - 1.03) |
| Temperature (annual average) | -0.04 | 0.88 (0.60 - 1.30) |
| Agricultural land use class (% of deciduous broadleaf forests) | -0.60 | 0.64 (0.05 - 2.77) |
| Agricultural land use class (% of mixed forests) | 0.05 | 1.03 (0.01 - 1.26) |
| Agricultural land use class (% of grasslands) | -0.24 | 0.80 (0.56 - 0.99) |

**Supplementary table 22. Associations of predator species richness with *Ebolavirus* incidence in Model 3, constructed using the species richness variables calculated with Maxent modeling results, and the only species reported to prey on bats were included.**

| **Variables** | **β (SE)** | **Odds ratio**  **(95 % CI)** |
| --- | --- | --- |
| Presence of order Strigiformes |  |  |
| without the predator | Reference | Reference |
| with the predator | 0.27 | 1.24 (0.56 - 3.24) |
| Presence of order Carnivora |  |  |
| without the predator | Reference | Reference |
| with the predator | -1.74 | 0.37 (0.01 - 1.89) |
| Presence of order Squamata |  |  |
| without the predator | Reference | Reference |
| with the predator | 1.36 | 4.01 (0.45 - 6.44) |
| Bat species richness (order Chiroptera) | -0.02 | 0.97 (0.93 - 1.15) |
| Human foot print score | 0.09 | 1.08 (0.97 - 1.25) |
| Precipitation (annual average) | 0.01 | 1.02 (0.98 - 1.03) |
| Temperature (annual average) | -0.16 | 0.88 (0.60 - 1.30) |
| Agricultural land use class (% of deciduous broadleaf forests) | -0.61 | 0.54 (0.01 - 1.07) |
| Agricultural land use class (% of mixed forests) | 0.03 | 1.03 (0.01 - 1.26) |
| Agricultural land use class (% of grasslands) | -0.20 | 0.82 (0.39 - 0.98) |

**Supplementary table 23. Associations of predator species richness with *Ebolavirus* incidence in Model 4, constructed using the species richness variables calculated with Maxent modeling results, and the only species reported to prey on bats were included.**

| **Variables** | **β (SE)** | **Odds ratio**  **(95 % CI)** |
| --- | --- | --- |
| Presence of order Strigiformes |  |  |
| without the predator | Reference | Reference |
| with the predator | 0.26 | 1.37 (0.84 - 2.81) |
| Presence of order Carnivora |  |  |
| without the predator | Reference | Reference |
| with the predator | -1.65 | 0.29 (0.01 - 1.32) |
| Presence of order Squamata |  |  |
| without the predator | Reference | Reference |
| with the predator | 1.11 | 5.13 (0.68 - 8.59) |
| Bat species richness (order Chiroptera) | -0.05 | 0.95 (0.86 - 1.03) |
| Human foot print score | 0.20 | 1.18 (1.01 - 1.31) |
| Precipitation (annual average) | 0.01 | 1.02 (0.98 - 1.03) |
| Temperature (annual average) | -0.04 | 0.88 (0.60 - 1.30) |
| Agricultural land use class (% of deciduous broadleaf forests) | -0.60 | 0.64 (0.05 - 2.77) |
| Agricultural land use class (% of mixed forests) | 0.05 | 1.03 (0.01 - 1.26) |
| Agricultural land use class (% of grasslands) | -0.24 | 0.80 (0.56 - 0.99) |

**Supplementary table 24. Associations of predator species richness with *Ebolavirus* incidence in Model 5, constructed using the species richness variables calculated with Maxent modeling results, and the only species reported to prey on bats were included.**

| **Variables** | **β (SE)** | **Odds ratio**  **(95 % CI)** |
| --- | --- | --- |
| Presence of order Strigiformes |  |  |
| without the predator | Reference | Reference |
| with the predator | 0.07 | 1.08 (0.43 - 2.51) |
| Presence of order Carnivora |  |  |
| without the predator | Reference | Reference |
| with the predator | -1.13 | 0.35 (0.05 - 1.74) |
| Presence of order Squamata |  |  |
| without the predator | Reference | Reference |
| with the predator | 1.06 | 4.08 (0.09 - 7.06) |
| Bat species richness (order Chiroptera) | 0.03 | 1.02 (0.93 - 1.12) |
| Human foot print score | 0.19 | 1.16 (1.01 - 1.31) |
| Precipitation (annual average) | 0.01 | 1.02 (0.98 - 1.03) |
| Temperature (annual average) | -0.12 | 0.88 (0.60 - 1.30) |
| Agricultural land use class (% of deciduous broadleaf forests) | -0.20 | 0.84 (0.33 - 1.49) |
| Agricultural land use class (% of mixed forests) | 0.01 | 1.03 (0.72 - 1.26) |
| Agricultural land use class (% of grasslands) | -0.24 | 0.60 (0.60 - 0.97) |

**Supplementary table 25. Associations of predator species richness with *Marburgvirus* incidence in fully adjusted model, using the species richness variables calculated with IUCN polygons.**

| **Variables** | **β (SE)** | **Odds ratio (95 % CI)** |
| --- | --- | --- |
| Species richness of order Strigiformes (no. species) |  |  |
| < 9 | Reference | Reference |
| 9 ≤ | 0.22 (0.39) | 1.25 (0.58 - 2.83) |
| Species richness of order Carnivora (no. species) |  |  |
| < 18 | Reference | Reference |
| 18 ≤ | 0.16 (0.38) | 1.18 (0.57 - 1.65) |
| Species richness of family Colubridae (no. species) |  |  |
| < 20 | Reference | Reference |
| 20 ≤ | 0.05 (0.16) | 1.05 (0.76 - 1.46) |

| Bat species richness (order Chiroptera) | 0.05 (0.06) | 1.05 (0.87 - 1.20) |
| --- | --- | --- |
| Human foot print score | -0.15 (0.11) | 0.86 (0.57 - 1.04) |
| Precipitation (annual average) | 0.01 (0.02) | 1.02 (0.94 - 1.08) |
| Temperature (annual average) | -0.32 (0.28) | 0.71 (0.33 - 1.39) |
| Gross Domestic Product (per capita) | 0.00 (0.00) | 1.00 (1.00 -1.00) |
| Population density (per km^2) | 0.01 (0.01) | 1.00 (0.98 - 1.01) |
| Agricultural land use class (% of savannas) | 0.01 (0.02) | 1.01 (0.99 - 1.02) |
| Agricultural land use class (% of grasslands) | -0.06 (0.07) | 0.94 (0.77 - 1.06) |
| Agricultural land use class (% of cropland/natural vegetation mosaics) | 0.08 (0.05) | 1.08 (0.99 - 1.22) |
| Agricultural land use class (% of deciduous broadleaf forests) | -0.10 (0.28) | 0.94 (0.47 - 1.38) |
| Agricultural land use class (% of mixed forests) | 0.08 (0.13) | 1.11 (0.82 - 1.32) |

**Supplementary table 26. Associations of predator species richness with *Marburgvirus* incidence in fully adjusted model, constructed using the species richness variables calculated with Maxent modeling results, and the only species reported to prey on bats were included.**

| **Variables** | **β (SE)** | **Odds ratio (95 % CI)** |
| --- | --- | --- |
| Species richness of order Strigiformes (no. species) |  |  |
| < 9 | Reference | Reference |
| 9 ≤ | 0.56 (0.58) | 1.77 (0.12 - 2.43) |
| Species richness of order Carnivora (no. species) |  |  |
| < 18 | Reference | Reference |
| 18 ≤ | -1.84 (0.77) | 0.13 (0.01 - 2.81) |
| Species richness of family Colubridae (no. species) |  |  |
| < 20 | Reference | Reference |
| 20 ≤ | -2.22 (0.60) | 0.07 (0.01 - 3.61) |

| Bat species richness (order Chiroptera) | -0.07 (0.16) | 0.93 (0.65 - 1.24) |
| --- | --- | --- |
| Human foot print score | -0.05 (0.16) | 1.09 (0.92 - 1.29) |
| Precipitation (annual average) | 0.01 (0.01) | 1.02 (0.98 - 1.03) |
| Temperature (annual average) | -0.32 (0.23) | 0.88 (0.60 - 1.30) |
| Gross Domestic Product (per capita) | 0.00 (0.00) | 1.00 (1.00 -1.00) |
| Population density (per km^2) | 0.01 (0.01) | 1.00 (0.98 - 1.01) |
| Agricultural land use class (% of savannas) | -0.30 (5.62) | 0.54 (0.01 - 1.07) |
| Agricultural land use class (% of grasslands) | -0.56 (4.14) | 1.03 (0.01 - 1.26) |
| Agricultural land use class (% of cropland/natural vegetation mosaics) | -0.02 (0.02) | 0.99 (0.96 - 1.02) |
| Agricultural land use class (% of deciduous broadleaf forests) | -0.06 (0.05) | 0.82 (0.39 - 0.98) |
| Agricultural land use class (% of mixed forests) | 0.03 (0.03) | 1.01 (0.96 - 1.10) |

**Supplementary figure 1. Variable collinearity heatmap.** Pearson correlation coefficient for a given pair of predictive variables in *Ebolavirus* model.

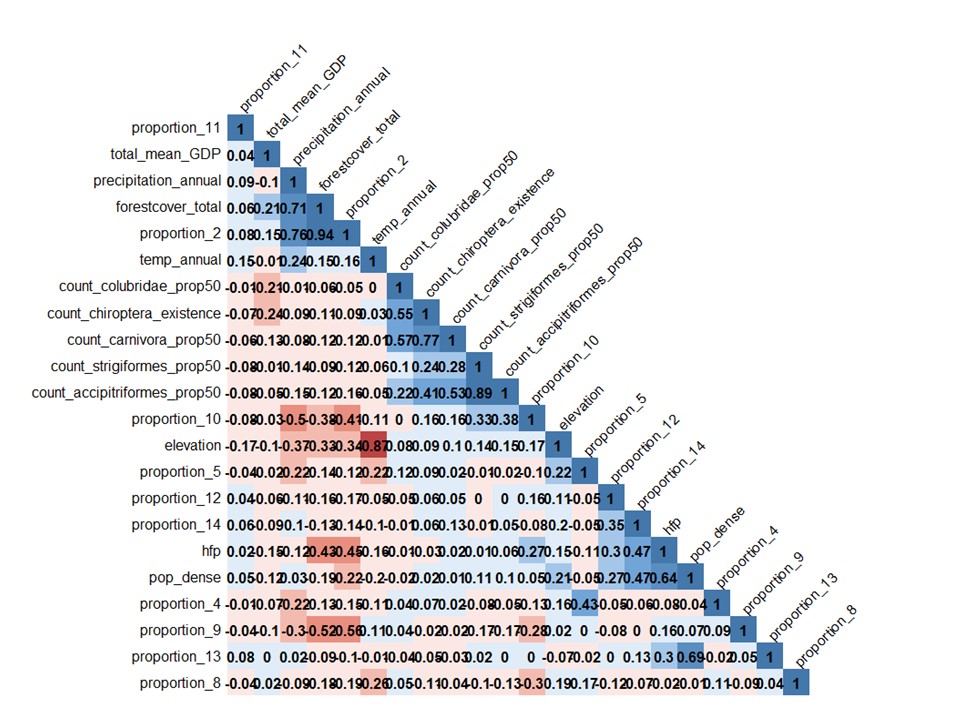

**Supplementary figure 2. Variable collinearity heatmap.** Pearson correlation coefficient for a given pair of predictive variables in *Marburgvirus* model.

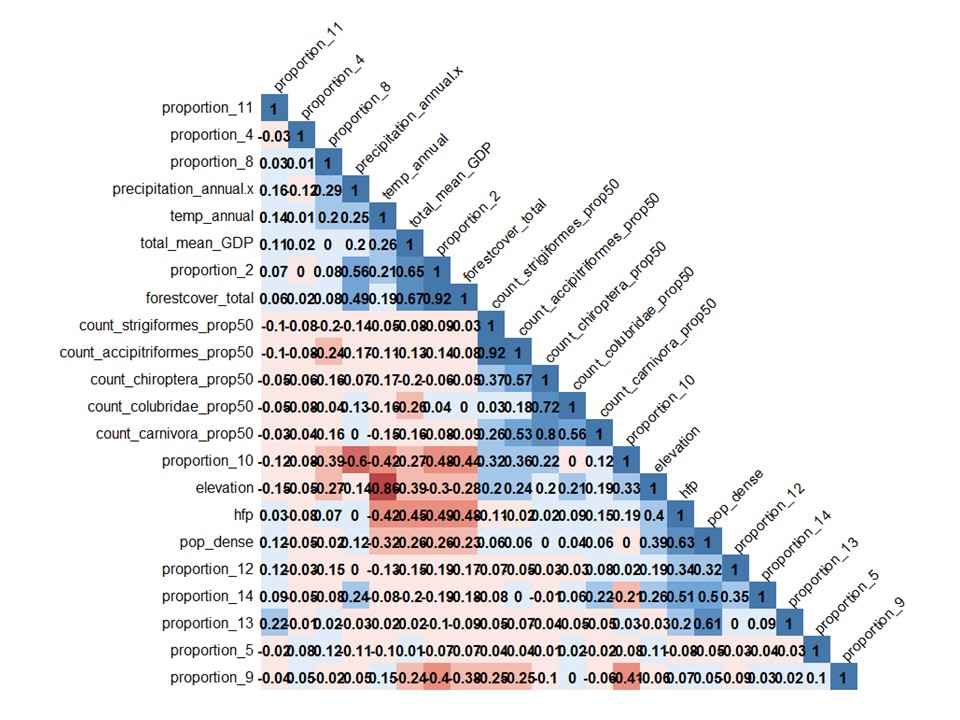

**Supplementary figure 3. Directed acyclic graph of the final models in the study.**

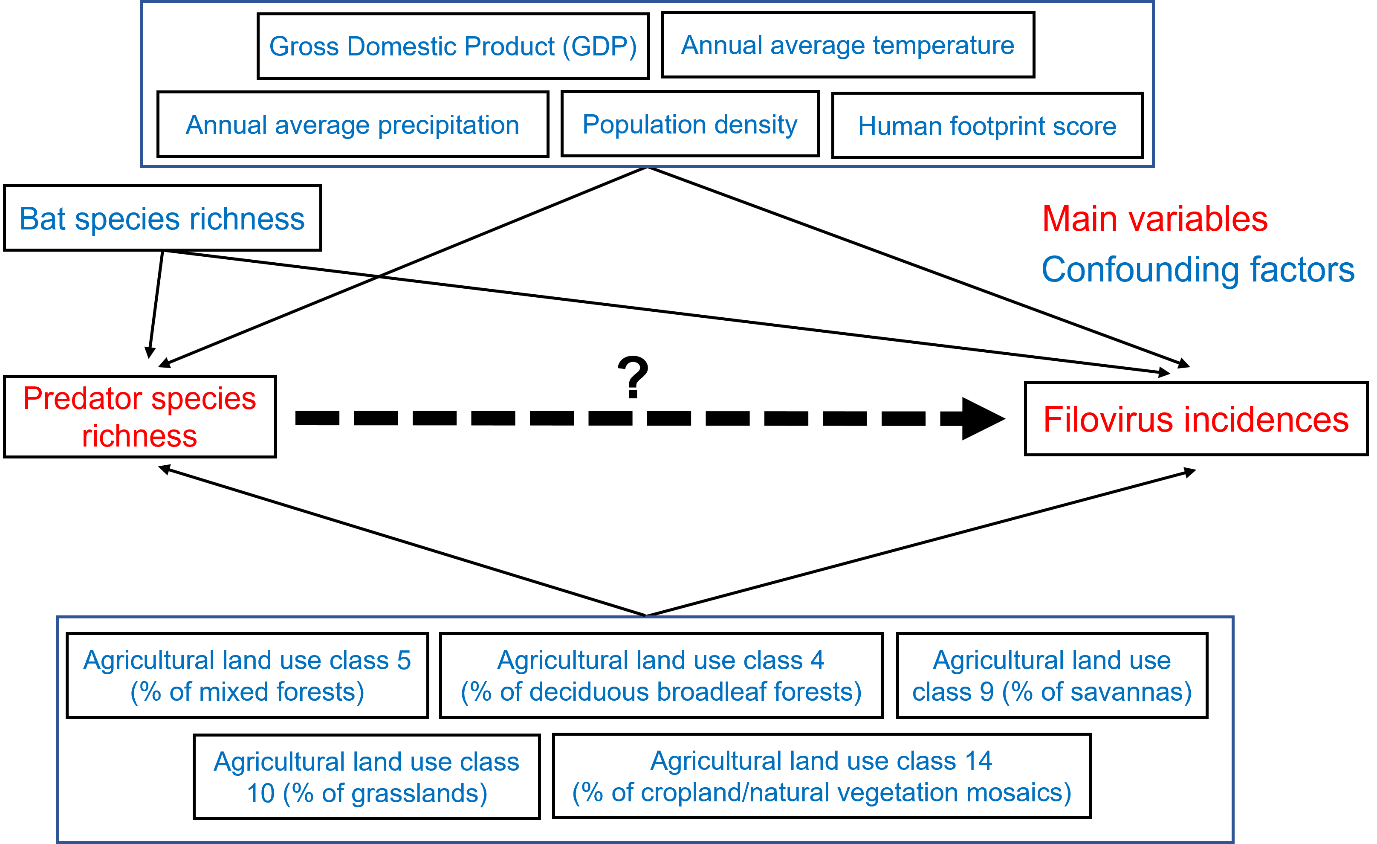

**Supplementary figure 4. Global Moran’s I plot and Moran’s I statistics for *Ebolavirus* incidence**

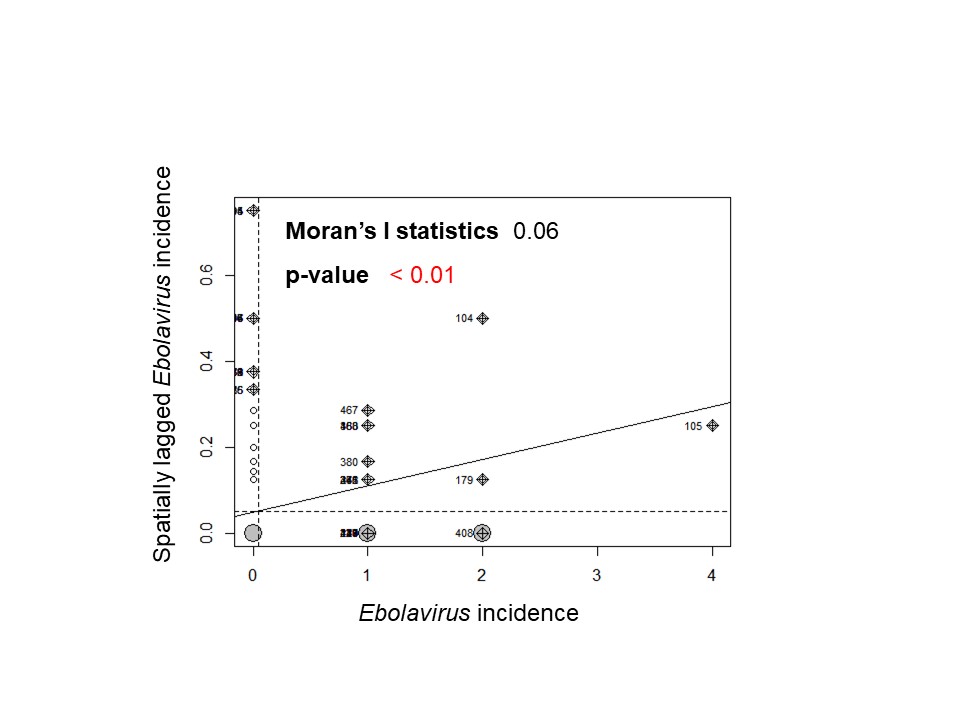

**Supplementary figure 5. Global Moran’s I plot and Moran’s I statistics for *Marburgirus* incidence**

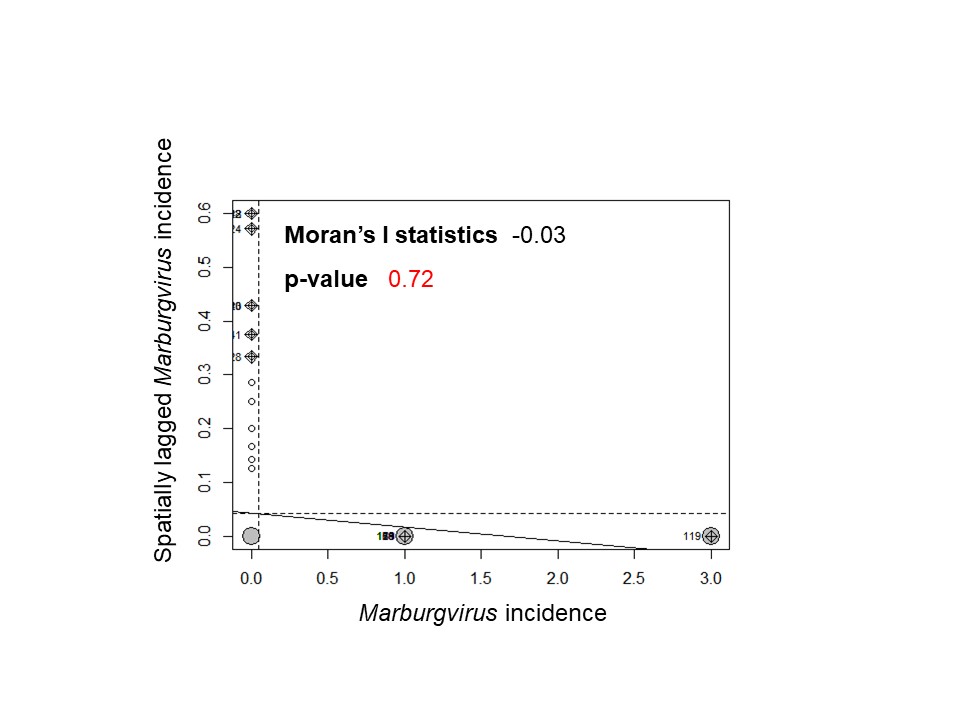

**Supplementary figure 6. Estimated ORs for *Ebolavirus* incidence according to the degree of species richness.** (A) The result of Model 1. (B) The result of Model 2. (C) The result of Model 3. (D) The result of Model 4. (E) The result of Model 5. The dots indicate the estimated ORs, with error bars representing the corresponding 95 % Wald’s credible intervals. Red means that the error bar does not intersect 1. The models were fitted with “R-INLA” package. The y-axis is shown on a logarithmic scale. The authors generated draws of each predator.

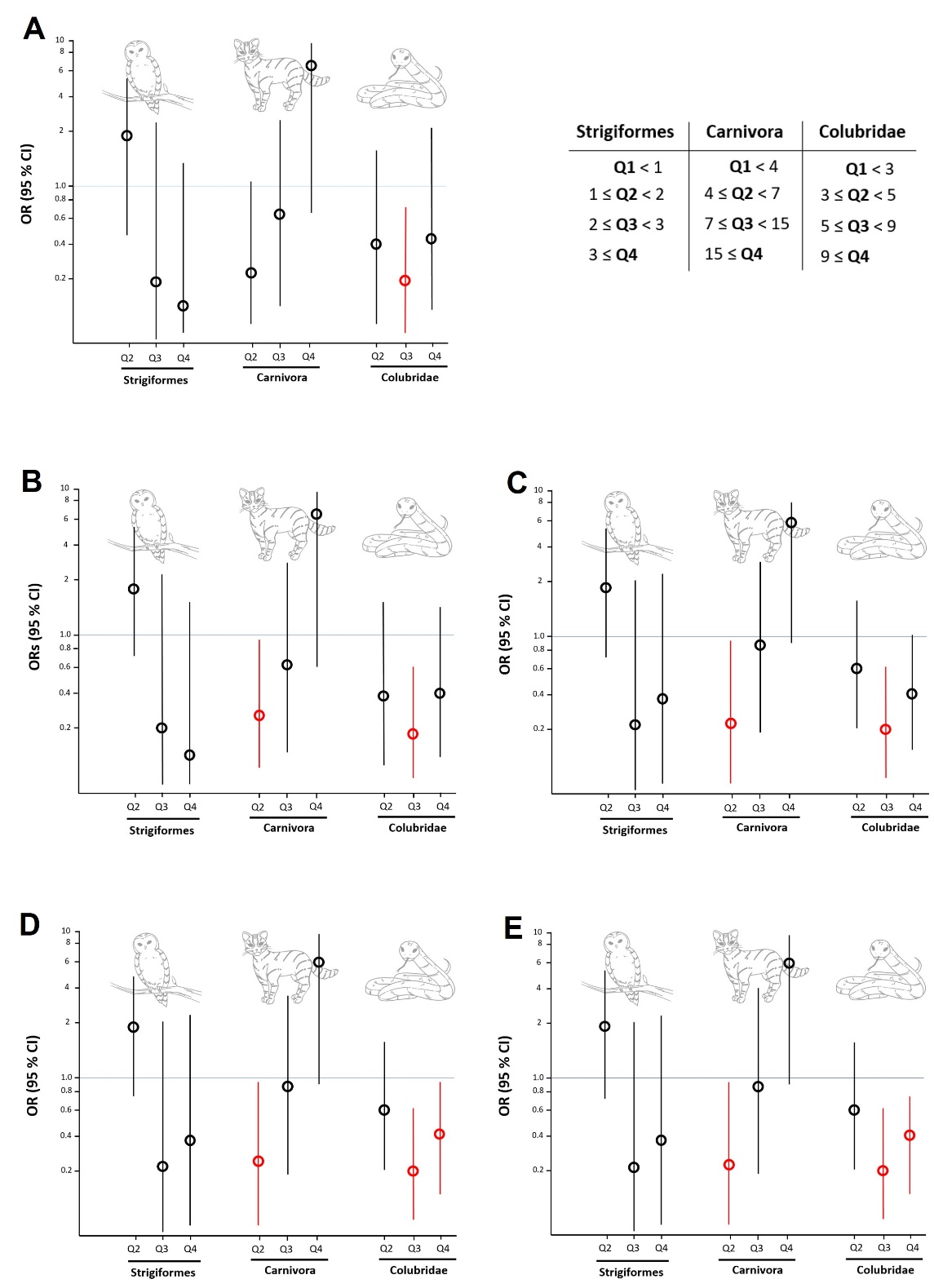

**Supplementary figure 7. Estimated ORs for *Ebolavirus* incidence according to the degree of species richness.** (A) The result of Model 1. (B) The result of Model 2. (C) The result of Model 3. (D) The result of Model 4. (E) The result of Model 5. The dots indicate the estimated ORs, with error bars representing the corresponding 95 % Wald’s credible intervals. Red means that the error bar does not intersect 1. The models were constructed using the species richness variables calculated with IUCN polygons. The y-axis is shown on a logarithmic scale. The authors generated draws of each predator.

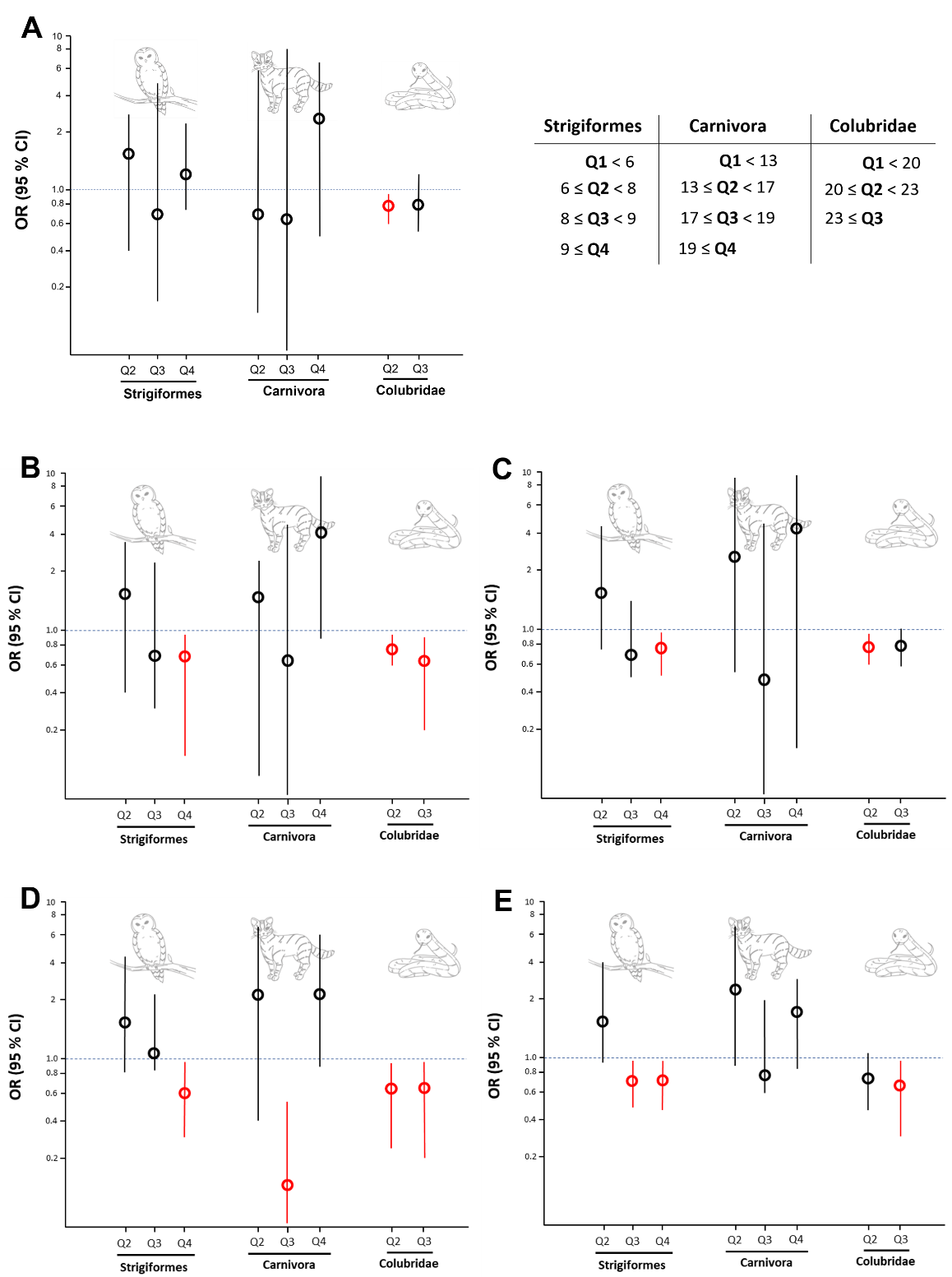

**Supplementary figure 8. Estimated ORs for *Ebolavirus* incidence according to the degree of species richness.** (A) The result of Model 1. (B) The result of Model 2. (C) The result of Model 3. (D) The result of Model 4. (E) The result of Model 5. The dots indicate the estimated ORs, with error bars representing the corresponding 95 % Wald’s credible intervals. Red means that the error bar does not intersect 1. The reference categories are when the species does not exist. The models were constructed using the species richness variables calculated with Maxent modeling results, and the only species reported to prey on bats were included. The y-axis is shown on a logarithmic scale. The authors generated draws of each predator.

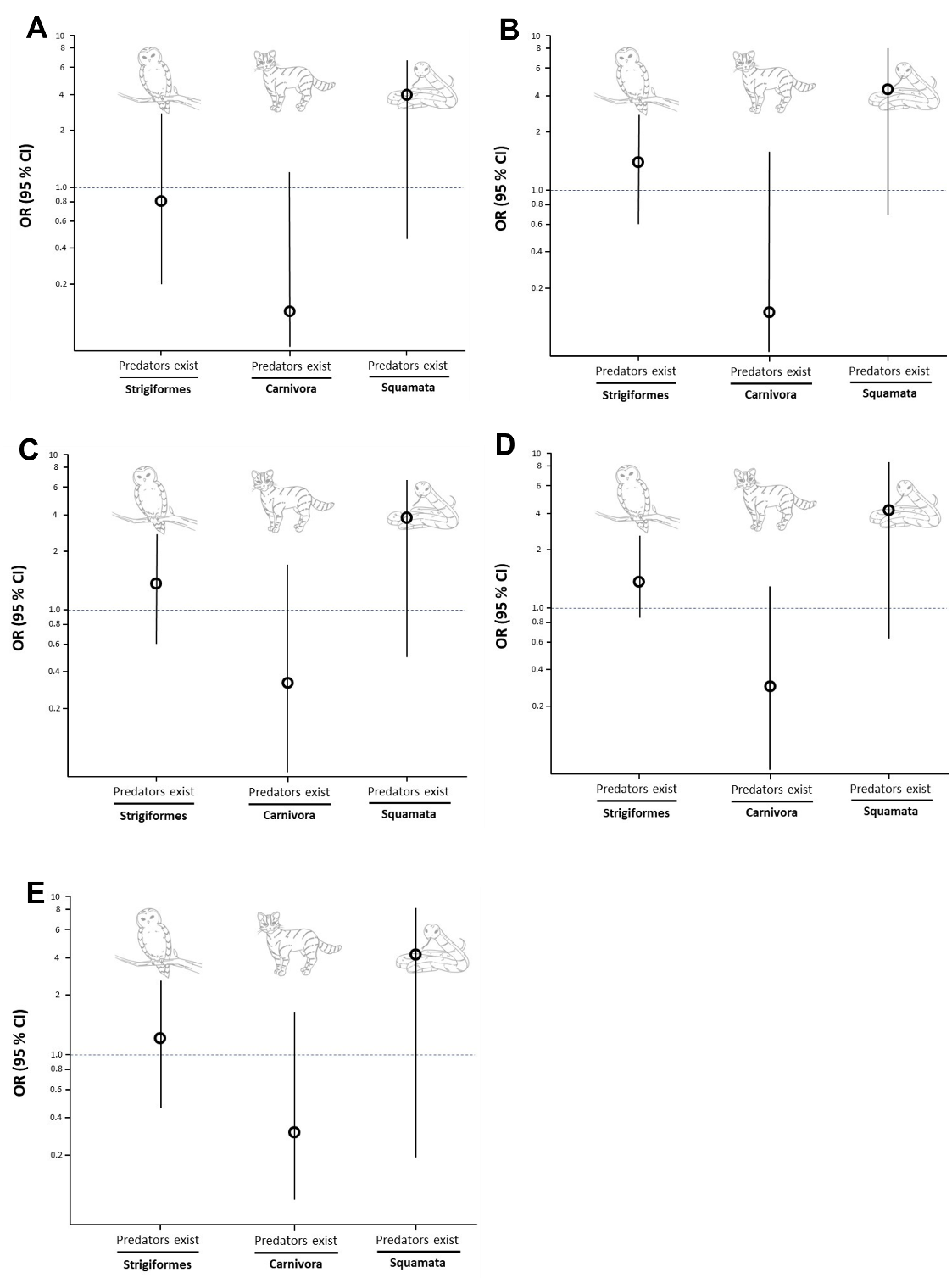

**Supplementary figure 9. Estimated ORs for *Marburgvirus* incidence according to the degree of species richness.** The dots indicate the estimated ORs, with error bars representing the corresponding 95 % Wald’s credible intervals. The model was constructed using the species richness variables calculated with IUCN polygons. The y-axis is shown on a logarithmic scale. The authors generated draws of each predator.

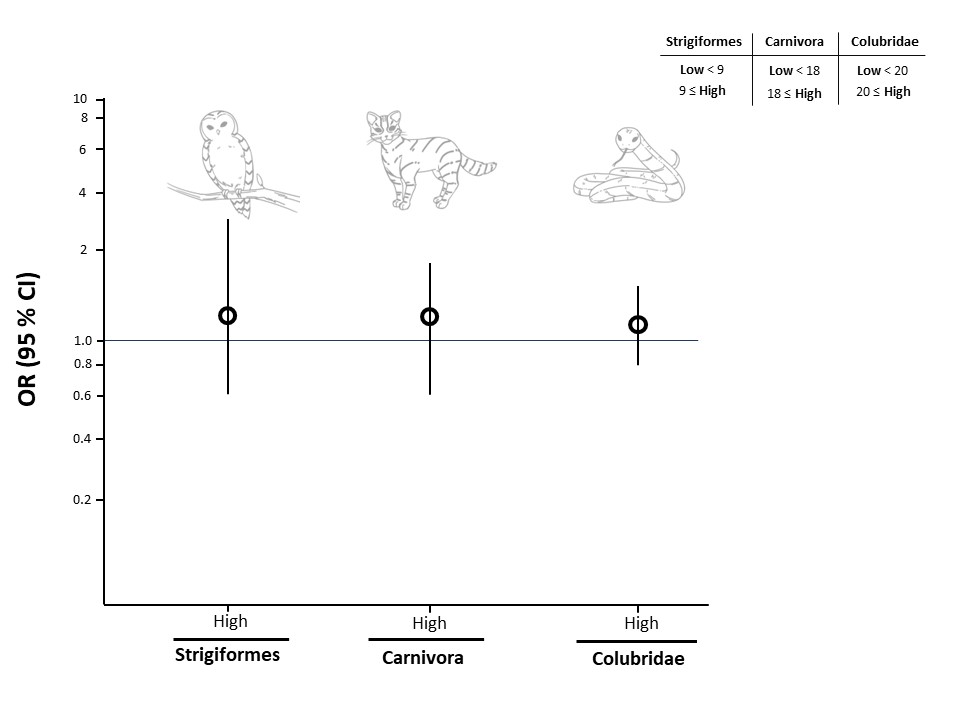

**Supplementary figure 10. Estimated ORs for *Marburgvirus* incidence according to the degree of species richness.** The dots indicate the estimated ORs, with error bars representing the corresponding 95 % Wald’s credible intervals. The reference categories are when the species does not exist. The models were constructed using the species richness variables calculated with Maxent modeling results, and the only species reported to prey on bats were included. The y-axis is shown on a logarithmic scale. The authors generated draws of each predator.

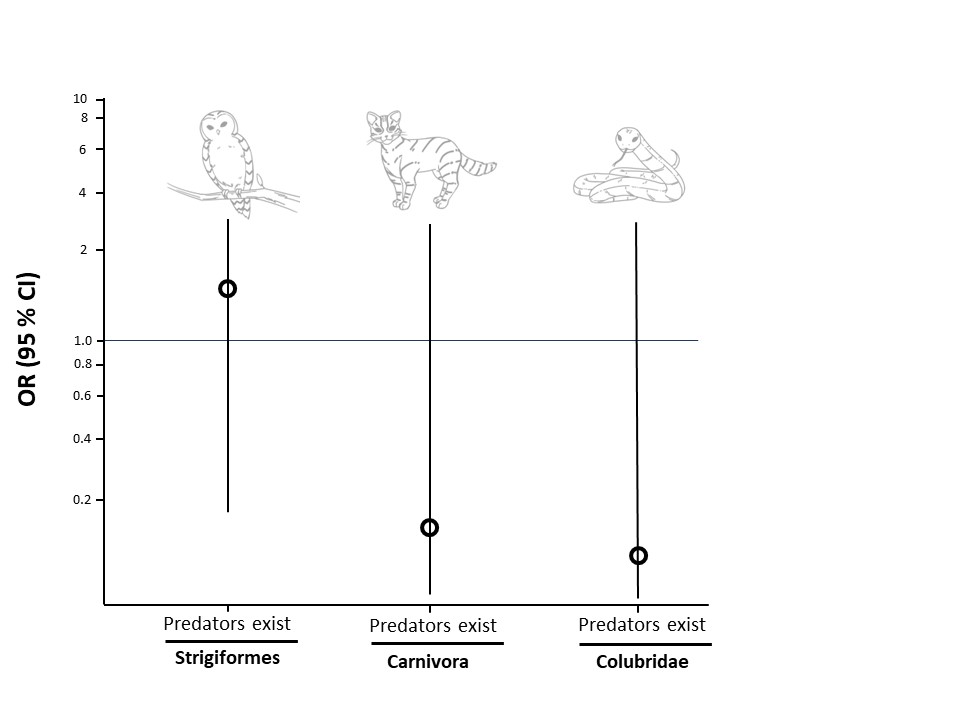
